# Powerful gene-based testing by integrating long-range chromatin interactions and knockoff genotypes

**DOI:** 10.1101/2021.07.14.21260405

**Authors:** Shiyang Ma, James L. Dalgleish, Justin Lee, Chen Wang, Linxi Liu, Richard Gill, Joseph D. Buxbaum, Wendy Chung, Hugues Aschard, Edwin K. Silverman, Michael H. Cho, Zihuai He, Iuliana Ionita-Laza

**Affiliations:** Department of Biostatistics, Columbia University, New York, NY, 10032, USA; Quantitative Sciences Unit, Department of Medicine, Stanford University, Stanford, CA, 94305, USA; Department of Statistics, University of Pittsburgh, Pittsburgh, PA, 15260, USA; Department of Human Genetics, Genentech, South San Francisco, CA, 94080, USA; Department of Epidemiology, Columbia University, New York, NY, 10032, USA; Departments of Psychiatry, Neuroscience, and Genetics and Genomic Sciences, Icahn School of Medicine at Mount Sinai, New York, NY, 10029, USA; Department of Pediatrics and Medicine, Herbert Irving Comprehensive Cancer Center, Columbia University Irving Medical Center, New York, NY, 10032, USA; Department of Computational Biology, Institut Pasteur, Paris, France; Channing Division of Network Medicine and Division of Pulmonary and Critical Care Medicine, Brigham and Women’s Hospital and Harvard Medical School, Boston, MA 02115, USA; Department of Neurology and Neurological Sciences, Stanford University, Stanford, CA 94305, USA

**Keywords:** gene-based association tests, long-range chromatin interactions, knockoff statistics, GWAS and whole-genome sequencing, fine-mapping

## Abstract

Gene-based tests are valuable techniques for identifying genetic factors in complex traits. Here we propose a novel gene-based testing framework that incorporates data on long-range chromatin interactions, several recent technical advances for region-based tests, and leverages the knockoff framework for synthetic genotype generation for improved gene discovery. Through simulations and applications to GWAS and whole-genome sequencing data for multiple diseases and traits we show that the proposed test increases the power over state-of-the-art gene-based tests in the literature, identifies genes that replicate in larger studies, and can provide a more narrow focus on the possible causal genes at a locus by reducing the confounding effect of linkage disequilibrium. Furthermore, our results show that incorporating genetic variation in distal regulatory elements tends to improve power over conventional tests. Results for UK Biobank and BioBank Japan traits are also available in a publicly accessible database that allows researchers to query gene-based results in an easy fashion.

**Significance:** Gene-based tests are important tools for elucidating the genetic basis of complex traits. Despite substantial recent efforts in this direction, the existing tests are still limited owing to low power and detection of false positive signals due to the confounding effects of linkage disequilibrium. In this paper we describe a novel gene-based test that attempts to address these limitations by incorporating data on long-range chromatin interactions, several recent technical advances for region-based testing, and the knockoff framework for synthetic genotype generation. Through extensive simulations and applications to multiple diseases and traits, we show that the proposed test increases the power over state-of-the-art gene-based tests and provides a narrower focus on the possible causal genes involved at a locus.

## 1 Introduction

Gene-based association tests are commonly used to identify genetic factors in complex traits. Relative to individual variant or window-based tests they have appealing features, including improved functional interpretation and potentially higher power due to lower penalty for multiple testing. Due to the recent advances in massively parallel sequencing technologies, a large number of gene-based tests have been proposed in the literature to test for association with genetic variation identified in sequencing studies^1–6^. One important limitation of the current gene-based tests is that they often fail to incorporate the epigenetic context in noncoding regions. Moreover, how to best analyze the noncoding part of the genome to increase power remains unclear. Recently, several sliding window approaches have been proposed to scan the genome with flexible window sizes, and appropriate adjustments for multiple testing while accounting for correlations among test statistics^7,8^. However, these approaches are essentially scanning the genome in a one-dimensional (1D) fashion and fail to take into account the three dimensional (3D) structure of the genome^9,10^. Furthermore, because they scan the genome agnostically, the burden of multiple testing is high which may lead to low power to identify true associations. These 1D approaches also suffer from interpretability issues similar to genomewide association studies (GWAS) and therefore require follow-up analyses to be performed in order to identify the target genes. Several existing tests such as MAGMA, H-MAGMA and STAAR-O^11–13^ attempt to link variants to their cognate genes based on physical proximity or chromatin interaction data. We will compare our proposed tests to these existing approaches both conceptually and empirically, and we will show that our tests are more flexible and powerful than these existing tests. Furthermore, when individual level data are available, the proposed tests can produce a more narrow list of associated genes at a locus by reducing the confounding effect of linkage disequilibrium (LD), a unique aspect of our gene-based test.

A related and popular gene-based strategy are the transcriptome-wide association studies (TWAS)^14,15^ that use GWAS data for a specific trait combined with genetic variation-gene expression repositories such as GTEx^16^ to perform gene-based association tests. However, TWAS are limited to eQTLs being present in the reference datasets, and the majority of genetic associations cannot be clearly assigned to existing eQTLs^17,18^. Therefore they may have reduced power to identify the relevant genes for the trait of interest.

Regulatory elements, including enhancers and promoters, play an important role in controlling when, where, and to what degree genes will be expressed. Most of the disease associated variants in GWAS lie in non-coding regions of the genome, and it is believed that a majority of causal noncoding variants reside in enhancers^19^. However, identifying enhancers and linking them to the genes they regulate is challenging. A number of methods have emerged in recent years to identify promoter-enhancer interactions. These techniques range from 3C (chromatin conformation capture), which is limited to the detection of a single interaction, to 4C (which can detect all loci that interact with a single locus), to many-to-many mapping technologies possible using targeted enrichment (e.g. ChIA-PET, HiChIP, and PLAC-seq). Hi-C maps the complete DNA interactome and elucidates the spatial organization of the human genome^20–23^. Hi-C provides direct physical evidence of interactions that may mediate gene regulatory relationships and can aid in identifying putative regulatory elements for a gene of interest. However, due to the prohibitive sequencing costs of the Hi-C experimental technique it is challenging to obtain high resolution (e.g. 1 Kb) Hi-C data in a large number of cell-types and tissues at multiple developmental times.

We propose here comprehensive gene-based association tests for common and rare genetic variation in both coding and noncoding, putative regulatory regions, and which incorporate several recent advances for region-based tests, including (1) scanning the genic and regulatory regions with varied window sizes, (2) the aggregated Cauchy association test to combine p-values from single variant, burden and dispersion (SKAT) tests, (3) incorporation of multiple functional annotations, and (4) the saddlepoint approximation for unbalanced case-control data^24–27^. To further improve the power and the ability to prioritize putative causal genes at significant loci when individual level data are available, we leverage a recent development in statistics, namely the knockoff framework for knockoff genotype generation^28^ that helps control the false discovery rate (FDR) under arbitrary correlation structure, and attenuates the confounding effect of LD. One can think of the knockoff genotypes as synthetic, noisy copies of the original genotypes which resemble the original data in terms of LD structure but are conditionally independent of the trait of interest, given the original genotypes. Although conventional methods such as the Benjamini-Hochberg procedure are also designed to control the FDR^29^, they cannot guarantee FDR control at the target level with arbitrarily correlated p-values. Furthermore, unlike the knockoff framework implemented here, the conventional methods do not naturally account for correlations due to LD.

We evaluate the performance relative to existing methods using simulations and applications to multiple studies, including GWAS studies for neuropsychiatric and neurodegenerative diseases, whole-genome sequencing studies for Alzheimer’s Disease from the Alzheimer’s Disease Sequencing Project (ADSP), and for lung function from the NHLBI Trans-Omics for Precision Medicine (TOPMed) Program. We also provide results of applications to UK Biobank and BioBank Japan binary and continuous traits.

## 2 Results

### 2.1 Overview of the proposed gene-based association tests

We provide here a brief overview of the proposed gene-based tests that aim to comprehensively evaluate the effects of common and rare, coding, proximal and distal regulatory variation on a trait of interest. A workflow depicting the overall gene-based testing approach proposed here is shown in Figure 1. Briefly, we build our final test, GeneScan3DKnock, progressively, starting with a test focused on scanning the gene body region (i.e. the interval between the transcription start site and the end of 3’ UTR) with varied window sizes. We refer to this test as GeneScan1D. We extend this test by incorporating genetic variants residing in putative regulatory elements such as promoters and enhancers. In particular, we use chromatin immunoprecipitation sequencing (ChIP-seq) peak data extracted from the ChIP-Atlas database to identify promoter regions, and data from the GeneHancer database to link enhancers to their target genes^30^. We also use the activity-by-contact (ABC) model to predict functional enhancer-gene connections for 5 cell types and tissues^31^. This is the GeneScan3D test. Finally, when individual level data are available, we implement the knockoff framework for a more powerful gene discovery and fine-mapping approach, and refer to this test as GeneScan3DKnock.

**Figure 1:**
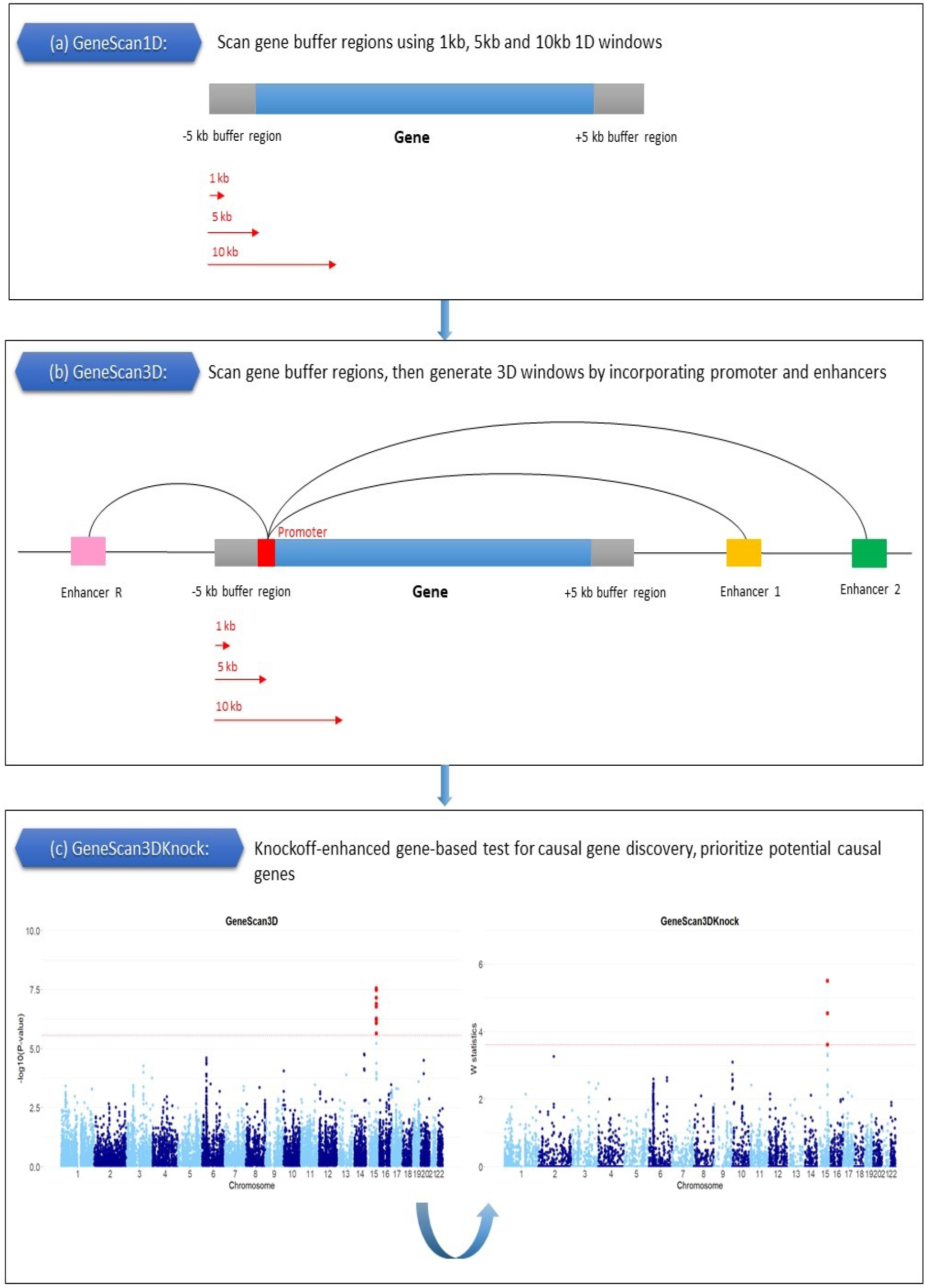
Workflow of the proposed gene-based tests. (a) GeneScan1D, a 1D scan of the gene and buffer region; (b) GeneScan3D, a 3D scan of the gene and regulatory elements linked to it; (c) GeneScan3DKnock, the knockoff-enhanced test, implementing a knockoff-based version of GeneScan3D.

We take advantage of recent advances in region-based tests for sequencing data^4,24,26^ to perform computationally efficient and comprehensive tests with genetic variation in a gene (including variants located in proximal and distal regulatory elements), while scanning the gene with a range of window sizes for improved power. The framework allows for the incorporation of a variety of functional genomics annotations as weights for individual variants included in the tests. Furthermore, a novel aspect of our testing framework is the derivation of knockoff statistics based on the generation of knockoff (synthetic) genetic data that resemble the original genotypes in terms of correlation structure but are conditionally independent of the outcome variable given the true genotypes^8,28,32^. The knockoff genotypes are essentially noisy copies of the original genotypes and serve as negative controls for the original genotype data; they help select important genes while controlling the FDR. GeneScan3DKnock computes for each gene a knockoff statistic W that measures the importance of each gene (similar to a p-value), and then uses the knockoff filter to detect genes that are significant at a specified FDR target level^28^. We also compute a q-value for each gene. A q-value is similar to a p-value, except that it measures significance in terms of FDR rather than FWER, and already incorporates correction for multiple testing. The knockoff version of the gene-based test has unique features relative to the standard gene-based tests, including improved power and ability to prioritize causal genes over associations due to LD. The details on these specific tests can be found in the Methods section.

We compare with the nearest competitor gene-based tests in the literature, namely MAGMA/H-MAGMA^11,12^, TWAS/FUSION^15^ and STAAR-O^13^. We also show comparisons with the recently proposed window-based test KnockoffScreen^8^.

### 2.2 Simulation studies

#### Power and Type-I error rate evaluation for a single gene

We conducted simulation studies in order to (1) examine the Type-I error rates of the proposed tests, GeneScan1D and GeneScan3D, under different significance levels and to (2) evaluate the potential power gain by incorporating the regulatory elements. For power comparisons we considered the nearest competitor gene-based tests, MAGMA/H-MAGMA and STAAR-O.

For Type-I error rate simulations, we use real COPDGene TOPMed whole-genome sequencing data, with *n* = 5, 593 for a continuous trait and *n* = 4, 450 for a binary trait. We randomly select 10 genes (average gene length 25 Kb) and for each selected gene, we randomly select *R* = 2 GeneHancer and ABC enhancers (average enhancer length 1.35 Kb). For each selected gene and the corresponding enhancers, we use the real genotype data, while the phenotype data are simulated as below:

- For a continuous trait: *Y*_*i*_ = *Z*_*i*_ + ϵ_*i*_,
- For a binary trait: logit(*P*(*Y*_*i*_ = 1| *Z*_*i*_)) = *α*_0_ + *Z*_*i*_,

where *Z*_*i*_ ∼ *N*(0, 1) is a covariate and ϵ_*i*_ ∼ *N*(0, 1) is the standard normal error; *Z*_*i*_ and ϵ_*i*_ are independent. For binary trait, equal number of cases and controls are simulated. For GeneScan1D and GeneScan3D we use two window sizes, 1 Kb and 5 Kb to scan the gene region. All variants, and common variants only are considered in the Type-I error rate simulation studies.

To evaluate power and compare with existing tests such as MAGMA/H-MAGMA and STAAR-O, we use the same whole-genome sequencing data. We randomly select 10 genes (average length 25 Kb) and, for each selected gene, we randomly select *R* = 10 GeneHancer and ABC enhancers (average enhancer length 1.87 Kb). Power is computed for each gene separately, and the average over the 10 genes is reported. We make use of the real genotypes for the selected genes plus and minus a 5 Kb buffer region, and for the corresponding enhancers. For each gene, the phenotype data are generated as follows:

- For a continuous trait: *Y*_*i*_ = *β*_1_*G*_*i*1_ + … *β*_*s*_*G*_*is*_ + *Z*_*i*_ + ϵ_*i*_,
- For a binary trait: logit(*P*(*Y*_*i*_ = 1| *Z*_*i*_, **G**_**i**_)) = *α*_0_ + *Z*_*i*_ + *β*_1_*G*_*i*1_ + … *β*_*s*_*G*_*is*_,

where *G*_*i j*_ denotes the genotypes of randomly selected causal variants and *β* _*j*_’s are the corresponding effect sizes. For binary trait, equal number of cases and controls are simulated. We set 2% of the variants in the gene and buffer region to be causal, all within a 2 Kb signal window. For each enhancer, we set 2% (uniformly distributed) variants to be causal. The effect size of the causal variant *j* is assumed to be *β* _*j*_ = *c* |log_10_MAF_*j*_|. We assume *c* = 0.25 for the continuous trait and *c* = 0.6 (e.g. OR = 6.05, when MAF = 0.001) for the binary trait.

For GeneScan1D and GeneScan3D we use three window sizes for scanning, 1 Kb, 5 Kb and 10 Kb. We apply MAGMA on the gene plus and minus 5 Kb buffer region. For GeneScan3D and H-MAGMA we incorporate *R* = {2, 5, 10} enhancers. We also conduct STAAR-O gene-centric analyses on (1) the entire gene body and (2) the same *R* = {2, 5, 10} enhancers, and then combine the STAAR-O p-values for these elements using the Cauchy combination method. As detailed in the Methods section, to allow for fair comparisons, for STAAR-O we use the same weighting and MAF/MAC thresholds as used for the proposed tests. For the sake of completeness, we also run the default setting of STAAR-O gene-centric analyses focused on rare variants. Finally, we adjust for 10 principal components of ancestry.

##### Type-I error rate

We conducted 10^7^ replications to examine the empirical Type-I error rate under both continuous and binary traits (Table S1). For continuous traits, the Type-I error rates were well controlled in all analyses under moderate significance levels 10^−3^, 10^−4^ and 10^−5^. Even for a stringent significance level of 2.5 × 10^−6^, the Type-I error rates fall within the 95% confidence interval: (1.52 × 10^−6^, 3.48 × 10^−6^). For binary phenotypes, the Type-I error rates are slightly conservative at different levels.

##### Power

We evaluated the empirical power at significance level 2.5 × 10^−6^ using 10,000 replications (Figures 2(a) and S1(a)). As shown, GeneScan3D and STAAR-O have important power advantages relative to H-MAGMA, likely due to their better tolerance of noisy variation, as also demonstrated below. GeneScan3D also exhibits higher power than STAAR-O, likely due to the sliding window scanning implemented in GeneScan3D. The 3D tests overall tend to be more powerful than the 1D tests, with the relative benefits diminishing as the number of signal enhancers decreases. STAAR-O with the default settings (focused on rare variants only) has lower power performance (Figure S2(a)) as expected given that our simulations include common causal variants, in addition to rare causal variants.

**Figure 2:**
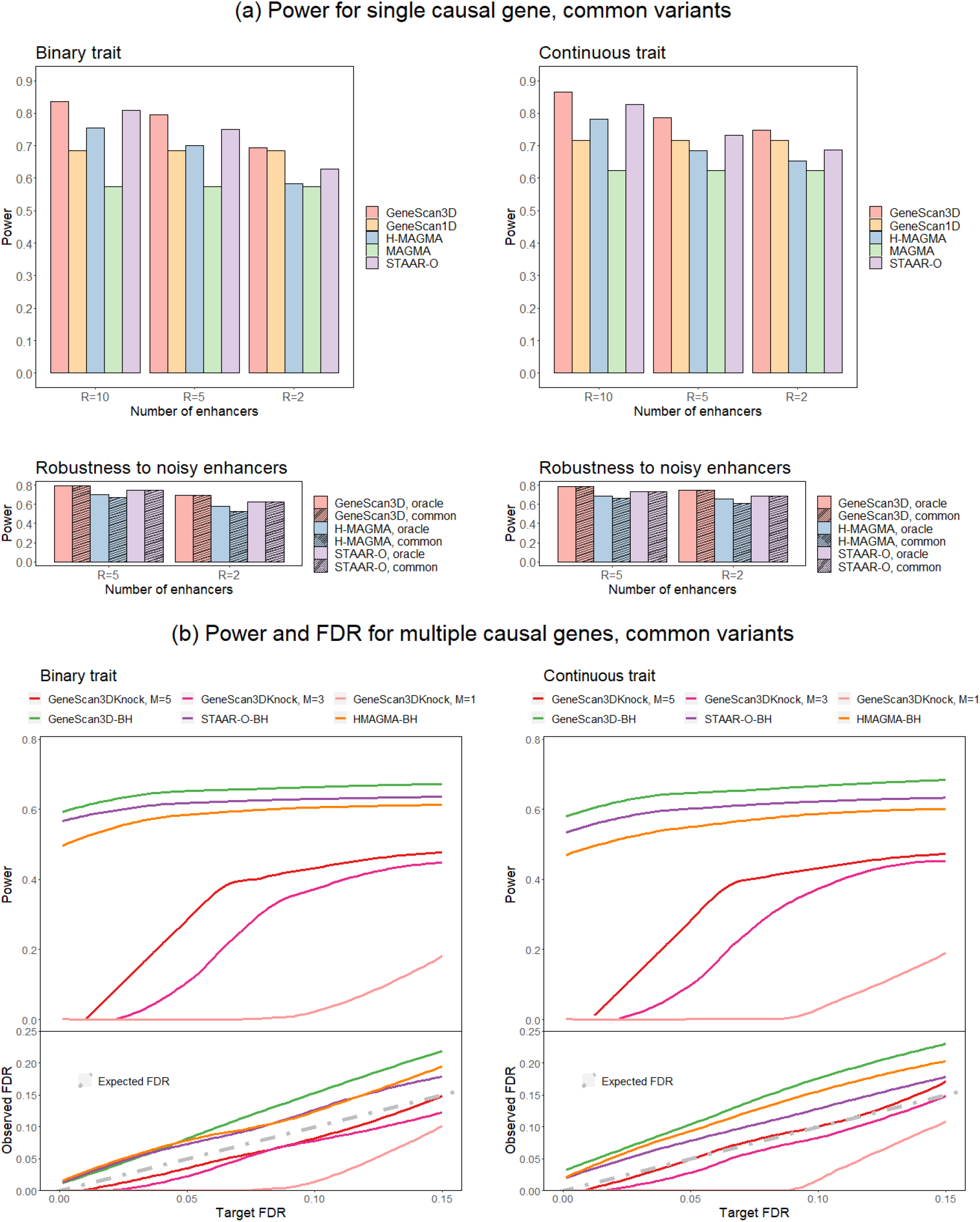
Power and false discovery rate of the proposed gene-based tests, binary and continuous traits with tests including only common variants. (a) Power and robustness to noisy enhancers. The top panels show power for the GeneScan3D, GeneScan1D, H-MAGMA, MAGMA and STAAR-O tests. The number of enhancers (*R*) ranges from 2 to 10. The bottom panels show power for the GeneScan3D, H-MAGMA and STAAR-O tests assuming causal variants in *R* = {2, 5} ‘causal’ enhancers. Power is compared between using only the *R* = {2, 5} ‘causal’ enhancers (the oracle approach) vs. using all 10 enhancers (including noisy enhancers). (b) Power and false discovery rate (FDR) for GeneScan3DKnock using different number of knockoffs and the Benjamini-Hochberg (BH) procedure for GeneScan3D, STAAR-O and H-MAGMA.

##### Robustness to noisy enhancers

When performing the 3D analyses, it is likely that some of the putative regulatory elements do not contain any signal variants. We conducted additional power simulation studies to evaluate the performance when only *R* = {2, 5} enhancers of a total of 10 enhancers for a gene contain any signal variants. We compared with the power of the oracle approach, i.e. when only the signal containing enhancers are included in the analyses (Figures 2(a), S1(a) and S2(a)). GeneScan3D and STAAR-O exhibit negligible power loss, suggesting that they are robust to inclusion of noisy enhancers, unlike H-MAGMA which is less robust in such realistic settings. This empirical observation is consistent with the theoretical expectation; while GeneScan3D/STAAR-O combine signal from individual enhancers using the Cauchy p-value combination method and hence are expected to maintain strong power in the presence of noisy enhancers, H-MAGMA is based on a principal component regression approach and hence combines genetic variants across multiple enhancers, rendering it less robust in the presence of noisy enhancers.

#### Power and false-discovery rate (FDR) evaluation with multiple causal genes

One novel aspect of the proposed knockoff-based test is that it allows for selecting significant genes by controlling the FDR in the presence of complex correlations due to LD. For the knockoff-based test, GeneScan3DKnock, we evaluate the empirical FDR and power assuming multiple causal and noise genes. We randomly select 10 causal genes, and 250 noise genes (gene length 10 Kb -100 Kb, average length 39 Kb) as follows. Among the noise genes, some are selected to be physically close to the causal genes, i.e. within ± 2 Mb region, and others are randomly selected across the genome. For each gene we only include the corresponding GeneHancer and ABC enhancers that fall within a 150 Kb region (± 75 Kb from the gene midpoint). This restriction to a 150 Kb region is done for computational reasons and only for these power simulations. On average, there are 10 enhancers for each gene, with an average length of 1.25 Kb. We generate multiple knockoff genotypes for 250-Kb regions spanning each gene (± 50 Kb on either side of the 150 Kb region) as detailed in the Methods section. Note that to avoid enhancer sharing across genes and too strong LD among causal and noise genes (which leads to false discoveries for all the statistical tests considered here) we select the genes such that the corresponding 150 kb regions are disjoint.

For each replicate, we randomly select a 10 Kb causal window in each causal gene plus and minus 5 Kb buffer region, and set 3.5% variants in the window to be causal. We also set 3.5% variants in all enhancers to be causal. We generate the continuous/binary traits using the selected causal variants as follows:

- For a continuous trait: *Y*_*i*_ = *Z*_*i*_ + *β*_1_*G*_*i*1_ + … + *β*_*s*_*G*_*is*_ + ϵ_*i*_,
- For a binary trait: logit(*P*(*Y*_*i*_ = 1| *Z*_*i*_, **G**_**i**_)) = *α*_0_ + *Z*_*i*_ + *β*_1_*G*_*i*1_ + … + *β*_*s*_*G*_*is*_.

As above, *Z*_*i*_ ∼ *N*(0, 1) is a covariate and ϵ_*i*_ ∼ *N*(0, 1) is the standard normal error; *Z*_*i*_ and ϵ_*i*_ are independent; *α*_0_ is chosen such that the prevalence is 10%. Again, we set the effect size *β* _*j*_ = *c* |log_10_MAF _*j*_| for the *j*-th causal variant, with *c* = 0.2 for the continuous trait and *c* = 0.6 for the binary trait.

The empirical power and FDR for GeneScan3DKnock are averaged over 100 replicates. We present results for single knockoff, as well as multiple knockoffs (*M* = 3 and 5). We calculate the original and knockoff p-values from the GeneScan3D test (for all variants and common variants), adjusting for 10 principal components of ancestry. We compute q-values for 10 causal and 250 noise genes in order to identify significant genes using the GeneScan3DKnock test at different target FDR levels, up to 0.15. The empirical power is defined as the proportion of causal genes being identified; the empirical FDR is defined as the proportion of detected genes that are noise. We show that GeneScan3DKnock can control the FDR at the target level, and using multiple knockoffs can improve power substantially, especially at lower levels of target FDR where the single knockoff approach has very low power as expected (Figures 2(b) and S1(b)).

For comparisons, we evaluate the empirical power and FDR for competitor methods including STAAR-O and H-MAGMA using the standard Benjamini-Hochberg (BH) procedure for FDR control. A gene is significant if the corresponding q-value ≤ the target FDR level. The results show that the conventional BH procedure may not control the FDR at the target level, and therefore the proposed knockoff-based approach provides a valid alternative when FDR control is desirable such as for polygenic traits with multiple underlying causal genes (Figures 2(b), S1(b) and S2(b)).

Although our main comparisons are with gene-based tests, we perform additional comparisons with the recently proposed window-based method KnockoffScreen^8^ in order to illustrate the need for a gene-based knockoff filter when our interest is controlling the FDR at the gene level. We apply KnockoffScreen by scanning each 150-Kb region using several window sizes (1 Kb, 5 Kb and 10 Kb) and we compute the empirical power and FDR of KnockoffScreen at both gene level and window level (Methods). Although KnockoffScreen can control the window-level FDR as shown in Figure S3, the empirical gene-level FDR can be quite high, suggesting that the proposed framework designed to control the FDR at the gene-level is more appropriate for gene discovery. Essentially, as a window-based test, KnockoffScreen leads to a larger number of rejections, i.e. higher power but also higher FDR at gene level (Figure S3).

### 2.3 Applications to individual level data from whole-genome sequencing studies

#### Alzheimer’s Disease

We present results from an application to whole-genome sequencing data from the Alzheimer’s Disease Sequencing Project (ADSP). The data include 3,085 whole genomes from the ADSP Discovery Extension Study and 809 whole genomes from the Alzheimer’s Disease Neuroimaging Initiative (ADNI), for a total of 3,894 whole genomes (more details are available in the Supplemental Material). We adjusted for age, age^2^, gender, ethnic group, sequencing center, and the leading 10 principal components of ancestry. Seven tissue/cell-type specific GenoNet functional scores^33^ related to brain are incorporated, including E071 (Brain hippocampus middle), E074 (Brain substantia nigra), E073 (Brain dorsolateral prefrontal cortex), E068 (Brain anterior caudate), E067 (Brain angular gyrus), E069 (Brain cingulate gyrus) and E072 (Brain inferior temporal lobe).

We show results for several tests including GeneScan1D and GeneScan3D tests, the proposed knockoff-based approach, GeneScan3DKnock, based on 5 random knockoffs, as well as existing tests including MAGMA/H-MAGMA, STAAR-O and TWAS. We do not include the results from KnockoffScreen since we have shown in the simulations that at the gene-level it can have inflated FDR. For all the tests except for the knockoff-based GeneScan3DKnock test we identify significant genes using the Bonferroni method for FWER control, since as shown in the simulations, the conventional Benjamini-Hochberg procedure does not control the FDR at the target level. For GeneScan3DKnock we use the implemented knockoff filter procedure to identify significant genes at an FDR threshold of 10%. We present results for common variants only (those with 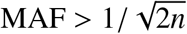 where *n* is the sample size), and all variants (rare variant only analyses are not well powered at these sample sizes).

Overall, all tests considered identify the well-known signal at the APOE locus (Table S2, Figures 3, and S4). The GeneScan3D, MAGMA-H and STAAR-O tests detect additional significant genes on chromosome 19, mostly due to signals residing in the promoters and/or enhancers overlapping genes at the *APOE* locus (Figure 4). These results suggest that the APOE region is a central nucleating point for loops that regulate expression of potentially Alzheimer’s Disease associated genes. Therefore, it is possible that the strong signal observed at the *APOE* locus can be linked to genes that are farther away (Figure 4 and Table S3).

**Figure 3:**
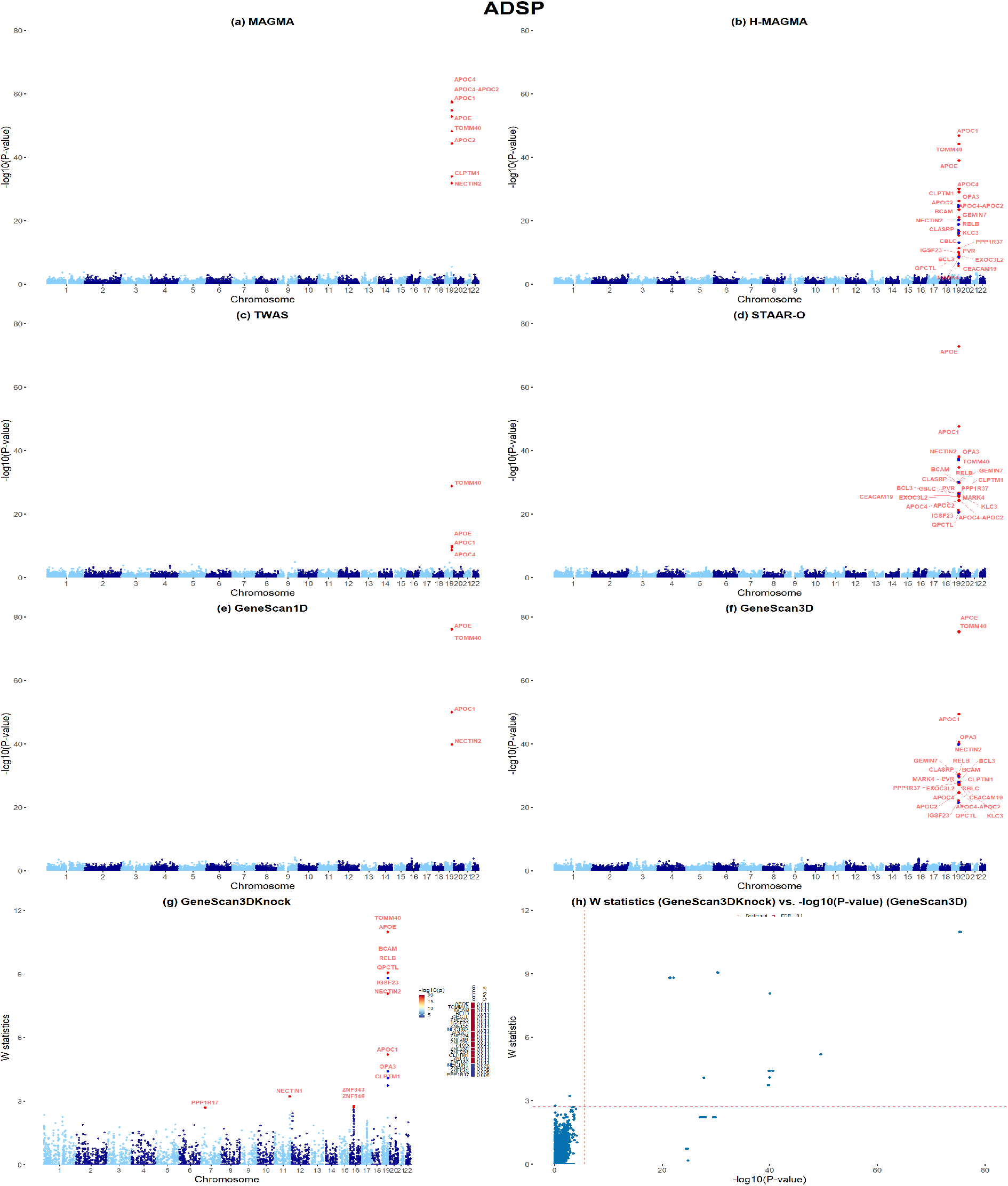
Applications to ADSP whole-genome sequencing data, common variants only. (a)-(f) Manhattan plots of MAGMA, H-MAGMA, TWAS, STAAR-O, GeneScan1D and GeneScan3D results, respectively. (g) Manhattan plot of GeneScan3DKnock results. Genes within the zinc-finger-containing (ZNF) gene cluster on chr19 are unlabeled and shown in blue in H-MAGMA, GeneScan3D and GeneScan3DKnock analyses for clear visualization. The right panel shows a heatmap with p-values of GeneScan3D test (truncated at 10^−20^) for all genes passing the FDR=0.1 threshold, and the corresponding q-values that already incorporate correction for multiple testing. The genes are shown in descending order of the knockoff statistics. (h) Scatter plot of *W* knockoff statistics (GeneScan3DKnock) vs. − log_10_(p value) (GeneScan3D) for common variants. Each dot represents a gene. The dashed lines show the significance threshold defined by Bonferroni correction (for p-values), and the data-adaptive threshold for false discovery rate control (FDR; for W statistic).

**Figure 4:**
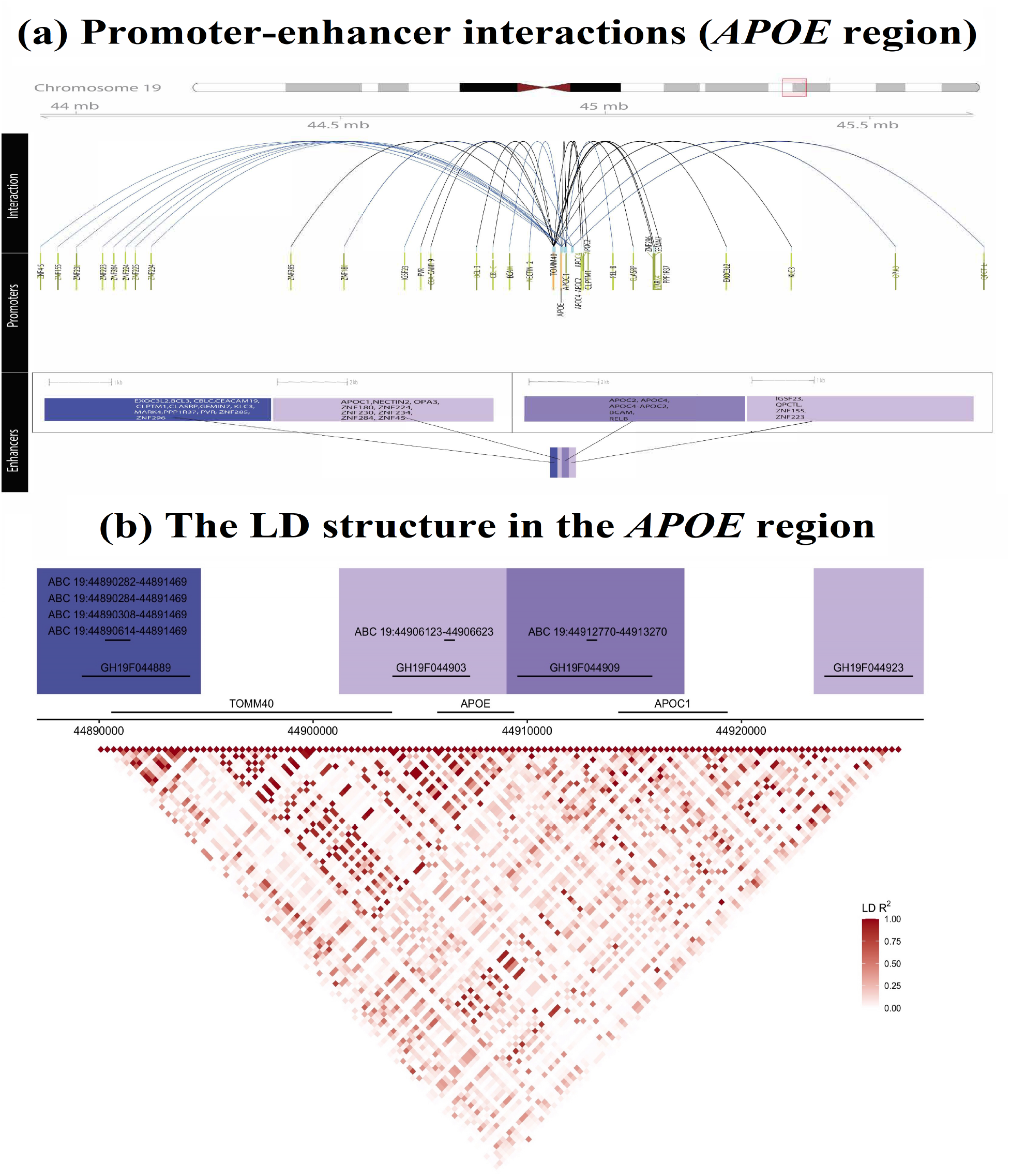
Visualization of promoter-enhancer interactions of significant genes at the *APOE* locus. (a) The promoter-enhancer links are shown for the significant genes in the GeneScan3D analyses for common variants, where arcs in blue point to those genes identified by the knockoff procedure only. Genes with a signal enhancer are shown in green; those with no signal enhancer in orange (middle panel). The *APOE* locus is the location of a tight cluster of several enhancers (shown in purple in the bottom panel), with arcs connecting the enhancers to many different gene promoters. (b) The LD structure in the *APOE* region. The locations of the enhancers in the region are also shown.

##### GeneScan3DKnock has improved power and reduces false-positive associations relative to existing tests

False positive signals can arise due to possible co-regulation of multiple genes by the same ‘causal’ enhancers, or simply due to LD among causal and non-causal variants in genes or associated regulatory elements. The knockoff-based test GeneScan3DKnock can not help eliminate false positives due to co-regulation, but can attenuate the effect of LD induced confounding. We compute the knockoff statistic *W* and q-value for each gene. A scatter plot of genome-wide W knockoff statistics vs. -log10(p-values) based on the GeneScan3D test illustrates how almost half of the significant genes at the *APOE* locus based on GeneScan3D are no longer significant in the GeneScan3DKnock test (Figures 3, S4, Tables S2, S4). These include a large number of genes linked to GH19F044889 and several overlapping ABC enhancers, that contain variants in high LD with variants in the *APOE* gene (Figure 4). Indeed, we obtained a narrower list of significant genes on chromosome 19 related to the *APOE* locus, that includes the main genes from the 1D tests, i.e. *APOE, TOMM40, APOC1* and *NECTIN2*, but other interesting genes as well, including *BCAM, RELB* and *QPCTL*. For example, rare variants in *BCAM* and *RELB* have recently been identified to be associated with AD and neuroimaging biomarkers of AD after adjusting for APOE genotypes^34,35^. *QPCT*, an important paralog of *QPCTL*, has been shown to be involved in AD pathogenesis and cognitive decline by glutaminyl cyclase (QC)-catalyzed pGlu-A*β* formation^36^. This ability to remove a substantial proportion of false positive signals due to LD is a unique and appealing feature of the proposed GeneScan3DKnock test.

Interestingly, GeneScan3DKnock detects several new associations outside chromosome 19 that are missed by the competitor gene-based tests. These include *NECTIN1, ZNF843, ZNF646, PPP1R17* (for common variants) and *HIPK3* (for all variants), which were previously found to be involved in AD-related pathophysiology. Nectin-1 is a member of the immunoglobulin superfamily and a Ca(2+)-independent adherens junction protein involved in synapse formation^37^. The important role of nectin in synaptic development and maintenance can explain how genetic variation in *NECTIN1* can perturb synaptic activity, and play a role in AD. *ZNF646* lies within the *KAT8* locus, recently identified in two large scale GWAS studies focused on clinically diagnosed AD and AD-by-proxy individuals^38,39^. Furthermore, *ZNF646* was prioritized at the *KAT8* locus based on high posterior probability for the colocalization between AD GWAS SNPs at the *KAT8* locus and eQTLs from both brain (dorsolateral prefrontal cortex) and microglia^40^. Similarly, *PPP1R17* was found to be significantly underexpressed in the brains of 14m old Sgo1 − /+ mice (a murine AD model of chromosome instability (CIN) with chromosomal and centrosomal cohesinopathy) compared with age matched wild type animals^41^. The protein encoded by this gene is found primarily in Purkinje cell bodies and projections in cerebellum and subsets of neurons in hypothalamus. A SNP located in the promoter of *PPP1R17* was previously found to be associated with hypercholesterolemia^42^. Finally, HIPK3 belongs to a group of homeodomain-interacting protein kinases (HIPKs), including HIPK2, which is downregulated by elevated amount of Amyloid *β* (A*β*), a hallmark of Alzheimer’s disease^43^. Protein HIPK3 levels were also found to be significantly different between individuals with mild cognitive impairment that converted to AD vs. the nonconverters^44^.

##### Replication of significant genes using summary statistics from a large meta-analysis AD study

To provide more objective evidence of replication, we leverage a large meta-analysis of clinical AD and AD-by-proxy studies (71,880 AD or proxy cases and 383,378 controls^56^) and perform gene-based tests (GeneScan1D and GeneScan3D) using the available GWAS summary statistics. Results are shown in Table S5. Note that most of the genes identified by GeneScan3DKnock at FDR 10%, including genes at the APOE locus, *ZNF646* and *PPP1R17*, have a replication p value based on GeneScan3D < 0.05, while genes identified by conventional BH controlling procedures (Figures S5-S6) fail to replicate for the most part, concordant with simulation studies showing that the BH procedure can result in inflated empirical FDR values and therefore it is not a rigorous procedure to identify significant genes at a desired FDR level.

#### Lung function (FEV1)

The Genetic Epidemiology of COPD (COPDGene) study includes chronic obstructive pulmonary disease (COPD) cases, controls, and additional smokers with varied lung function. In addition to COPD case/control status, lung function measurements are also available, including forced expiratory volume in one second (FEV1), forced vital capacity (FVC) and their ratio (FEV1/FVC). We analyzed whole genome sequencing data from the TOPMed freeze5b dataset, which includes a subset of 5,593 NHW individuals for continuous traits and 4,450 individuals for COPD case/control binary trait. We present results from the application to FEV1, adjusting for sequencing center, 10 principal components of ancestry, age, age^2^, gender, height, height^2^, smoking pack-years, and current smoking. We incorporated five tissue/cell-type specific GenoNet functional scores^33^ related to lung, namely: E017 (IMR90), E088 (Fetal lung), E096 (Lung), E114 (A549) and E128 (NHLF lung fibroblast).

As with the AD example, GeneScan3DKnock identifies new significant genes that are missed by the other tests. Specifically, we identify a new cluster of significant genes on chromosome 12 that includes *FRS2, CCT2, RAB3IP, LRRC10, BEST3* and that is missed by all the other gene-based tests considered (Figure 5). Notably, an intronic SNP (rs10444582) in *FRS2* was identified to be significantly associated with FEV1 in the UK Biobank and SpiroMeta (*p* = 1.2 × 10^−10^, *n* = 396, 723)^45^. This locus was not included in the final list of loci released by Shrine et al.^45^ as the p-value in the replication cohort (SpiroMeta) was only 3.5 × 10^−3^, above the predetermined significance threshold. As COPDGene was not part of the Shrine et al. study^45^, our findings on chromosome 12 provide additional, independent evidence for this signal. Another significant and potentially interesting gene is *RAB7A*. A common SNP (rs9847178) residing in the promoter flanking region of *RAB7A* has been found genome-wide significant in a recent large GWAS study on smoking^46^. A nearby SNP (rs7650872) in the same promoter flanking region has been found genome-wide significant with eosinophil counts in the UK Biobank^47^. High eosinophil counts predict decline in FEV1^48^. Interestingly, a recent study has showed that loss of *RAB7A* confers resistance to severe acute respiratory syndrome coronavirus 2 (SARS-CoV-2) by reducing ACE2 levels^49^, concordant with reports in the literature of loci associated with susceptibility and/or response to infection that have been previously associated with lung function phenotypes^50^.

**Figure 5:**
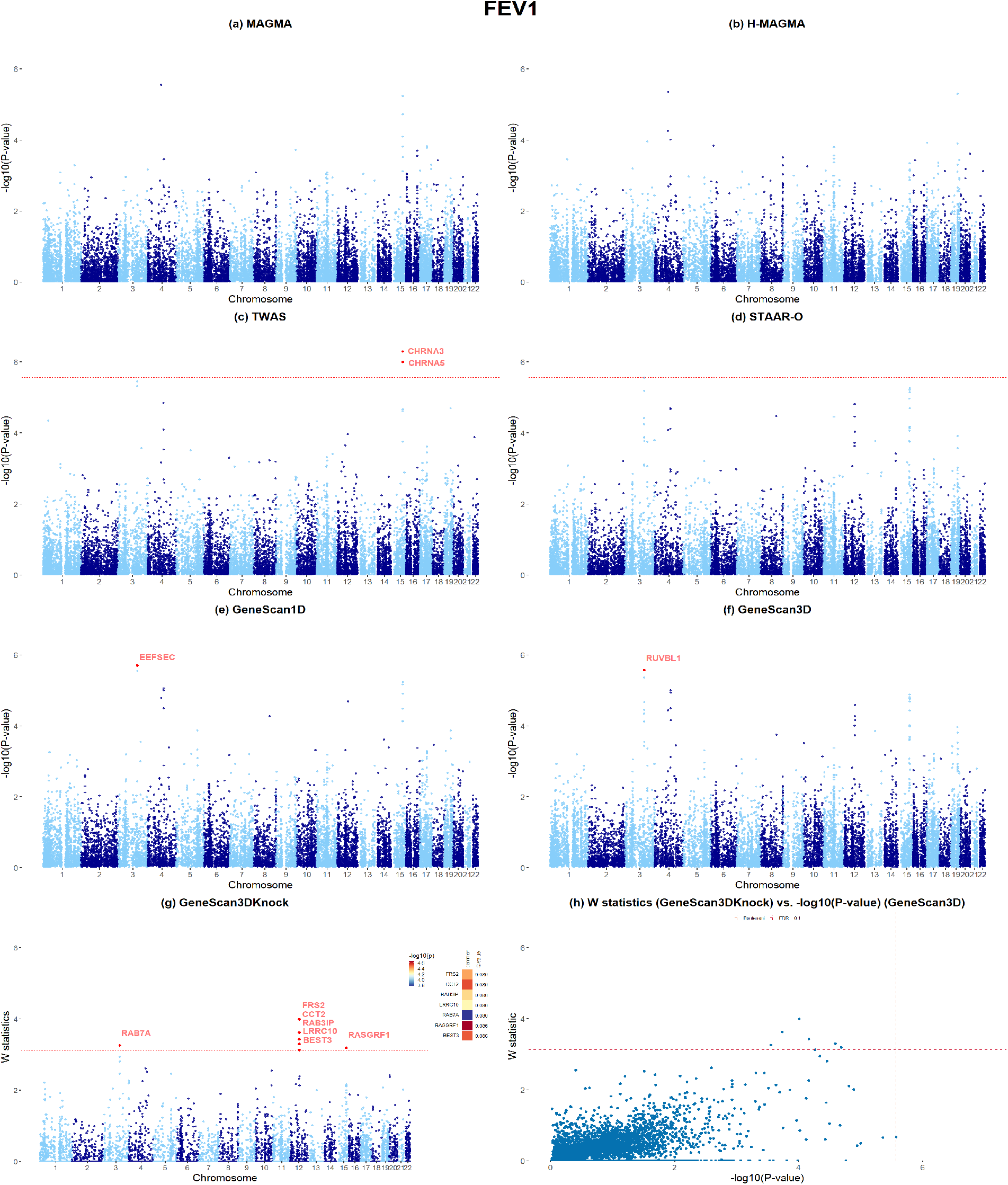
Applications to COPDGene whole-genome sequencing data (trait FEV1), for common variants only. (a)-(f) Manhattan plots of MAGMA, H-MAGMA, TWAS, STAAR-O, GeneScan1D and GeneScan3D results, respectively. (g) Manhattan plot of GeneScan3DKnock results. The right panel shows a heatmap with p-values of GeneScan3D test for all genes passing the FDR=0.1 threshold, and the corresponding q-values that already incorporate correction for multiple testing. The genes are shown in descending order of the knockoff statistics. (h) Scatter plot of *W* knockoff statistics (GeneScan3DKnock) vs. − log_10_(p value) (GeneScan3D) for common variants. Each dot represents a gene. The dashed lines show the significance threshold defined by Bonferroni correction (for p-values), and the data-adaptive threshold for false discovery rate control (FDR; for W statistic).

##### Replication of significant genes using summary statistics from the UK Biobank data

We performed similar replication studies for significant genes identified for FEV1 using 383,471 European individuals with FEV1 measurements in the UK biobank (Table S6). The covariates adjusted for in the analyses include 10 principal components of ancestry, age, age^2^, gender, age ·gender, and age^2^ ·gender. Note that the number of available covariates in the UK Biobank is limited, and some important covariates for FEV1 such as height and smoking are not adjusted for in these analyses. Despite this caveat, most of the genes identified as significantly associated with FEV1 in our COPDGene study replicate in the UK Biobank study.

### 2.4 Applications to GWAS summary statistics

#### GWAS for neuropsychiatric and neurodegenerative diseases

We apply the different gene-based tests to summary statistics from nine GWAS studies of brain disorders, including five neuropsychiatric traits: attention-deficit/hyperactivity disorder (ADHD; 20,183 cases and 35,191 controls)^51^, autism spectrum disorder (ASD; 18,381 cases and 27,969 controls)^52^, bipolar disorder (BD; 20,352 cases and 31,358 controls)^54^, schizophrenia (SCZ; 40,675 cases and 64,643 controls)^53^ and major depressive disorder (MDD; 170,756 cases and 329,443 controls)^55^; and four neurodegenerative traits: Alzheimer’s disease (AD; 71,880 cases and 383,378 controls)^56^, Parkinson’s disease (PD; 33,674 cases and 449,056 controls)^57^, amyotrophic lateral sclerosis (ALS; 12,577 cases and 23,475 controls)^58^ and multiple sclerosis (MS; 4,888 cases and 10,395 controls)^59^. We do not include STAAR-O here since the current implementation is not applicable to summary statistics.

Since these applications focus on brain disorders, we leverage two existing Hi-C human brain datasets for the dorsolateral prefrontal cortex (DLPFC) in adult brain^60^ and for the germinal zone (GZ) and cortical and subcortical plate (CP) in fetal brain^61^, and use the Fit-Hi-C method to identify statistically significant promoter-enhancer interactions from these data^62,63^ (Methods). Similarly, we apply MAGMA/H-MAGMA^12^ to the same datasets using the same significant Hi-C interactions. We also apply TWAS/FUSION^15^ based on 13 brain regions from GTEx v7 (Amygdala, Anterior cingulate cortex, Caudate, Cerebellar Hemisphere, Cerebellum, Cortex, Frontal Cortex, Hippocampus, Hypothalamus, Nucleus accumbens, Putamen, Spinal cord and Substantia nigra). The Cauchy p value combination method is used to combine TWAS p-values from different brain regions.

We use a liberal significance threshold (10^−3^) to select genes from these analyses (because some of the GWAS studies, e.g. AD, ADHS, MS, are underpowered) and investigate their expression patterns using spatiotemporal and single-cell transcriptomics data as described below. The number of significant genes at this threshold and the overlap across the different tests are shown in Table S7 and Figure S11. Compared with 1D analyses (GeneScan1D and MAGMA), GeneScan3D and H-MAGMA detect a much larger number of disease-associated genes, as expected given they incorporate signal from distal regulatory elements. GeneScan3D and H-MAGMA also detect a substantially higher number of significant genes relative to TWAS, a possible reflection of the limitation of eQTL-based approaches to discover significant associations that cannot be explained by eQTLs in the reference datasets.

##### Developmental and single cell expression profiles

We use spatiotemporal transcriptomic data from embryonic and adult brains measured at 15 time periods, ranging from 4 postconceptional weeks (PCW) to age ≥ 60 years^64^. The gene expression data are available for 6 brain regions: neocortex (NCX), mediodorsal nucleus of the thalamus (MD), cerebellar cortex (CBC), hippocampus (HIP), amygdala (AMY), and striatum (STR). We focus here on the cortical expression profiles (NCX area), with 410 samples for the prenatal stages (periods 1-7) and 526 samples for the postnatal stages (periods 8-15). We center the developmental expression matrix to the mean expression level for each sample.

We compute the average expression values across the significant genes for each brain sample, and then compare the values for prenatal and postnatal brain samples. For psychiatric diseases, the genes detected by GeneScan3D tend to have significantly higher expression in prenatal relative to postnatal periods as expected, and the trajectories highlight developmental windows in early or mid-gestation periods (Figure 6(a)). For neurodegenerative diseases, the pattern is reversed, with higher expression in the postnatal periods (except for MS), concordant with the expectation that genes for neurodegenerative disorders have increased expression with aging. The results for H-MAGMA suggest similar patterns, but with reversed patterns for ASD and ALS, and less significant differences for AD (Figure S12(a)). Results for GeneScan1D, MAGMA and TWAS show similar patterns (Figures S13-S15), although there are some discrepancies including the higher postnatal expression vs. prenatal expression for the ASD significant genes and significantly higher prenatal expression for ALS significant genes (MAGMA).

**Figure 6:**
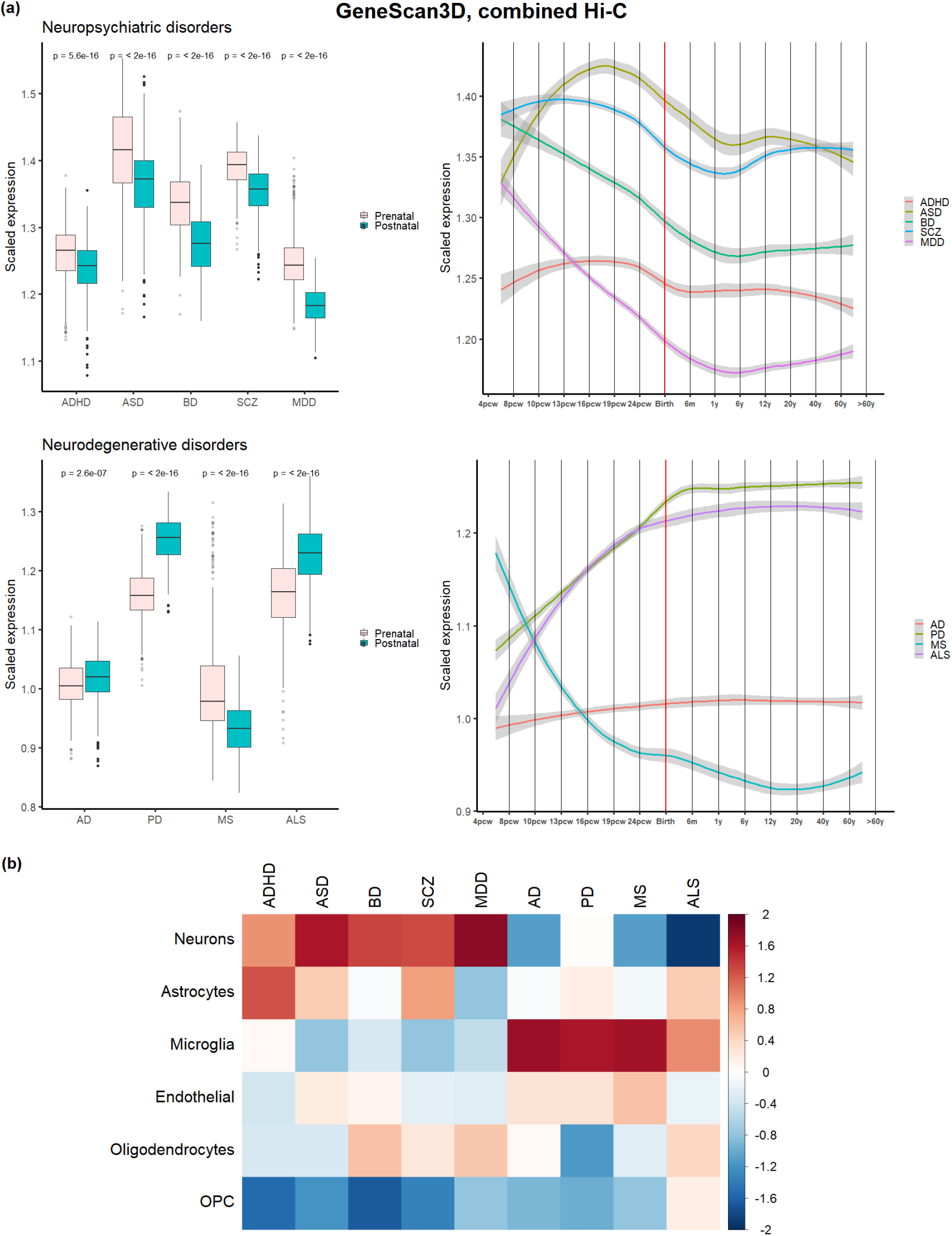
**(a) Human brain developmental expression of GeneScan3D significant genes for each brain-disorder**, combined Hi-C for Adult brain, Fetal brain GZ and CP layers. P-values of Wilcoxon rank sum tests are shown in the boxplots to compare independent prenatal and postnatal samples. **(b) Cell-type expression profiles of GeneScan3D significant genes**.

Additionally, we also leverage existing single-cell expression profiles^60^ on 285 single cells from 6 adult brain cell-types including neurons (131 cells), astrocytes (62 cells), microglia (16 cells), endothelial (20 cells), oligodendrocytes (38 cells) and oligodendrocyte progenitor cells (OPC, 18 cells). For each single cell, we center the expression data to the mean level of genes and then compute the average across the significant genes for a given disease. For each specific cell-type, we average across the multiple cells in this cell type. We compute standardized expression levels (i.e., subtract the mean and divide by the standard deviation) for the 6 adult cell-types. Genes identified by GeneScan3D for psychiatric disorders tend to show higher expression levels primarily in neurons, and to some extent in astrocytes compared to other cell types, whereas genes for neurodegenerative disorders tend to show higher expression levels primarily in microglia (Figure 6(b)). In particular, genes significant for ADHD show the highest expression in astrocytes, consistent with recent evidence suggesting a key role of astrocytes in the regulation of attention deficit disorder and hyperactivity^65^.

Results for the other tests show similar overall patterns as the GeneScan3D (Figures S12-S15), although with some differences, including less pronounced evidence for the role of astrocytes in ADHD (except for TWAS). These results serve as a proof of concept for the proposed 3D test, showing that genes identified by GeneScan3D and other existing tests exhibit expression patterns consistent with existing literature, i.e. an important role for neurons for neuropsychiatric diseases, and microglia for neurodegenerative diseases^66^.

#### Browser for results on UK/Japan Biobank data

##### UK Biobank

We have applied GeneScan3D to 1,403 UK Biobank binary phecodes and 827 continuous phenotypes using summary statistics on 28 million imputed variants. We have created a browser that displays phenome-wide results for a given gene, and genome-wide gene-based results for a given trait, and provides summary tables for significant genes. These gene-based results for the UK Biobank traits complement existing databases for single-variant tests^67^, and rare variant focused tests such as SAIGE-GENE^6^.

##### BioBank Japan

For non-European populations, we applied GeneScan3D to BioBank Japan binary phenotypes using available case-control GWAS summary statistics on 8,712,794 autosomal variants and 207,198 X chromosome variants, with 212,453 Japanese individuals across 42 diseases^68^. Results can be queried using the aforementioned browser.

## 3 Discussion

We propose novel gene-based tests that integrate genetic variation residing in putative regulatory regions and implement the knockoff framework for increased power and improved causal gene prioritization. This framework provides a rich toolkit for the analysis of GWAS and whole-genome sequencing data with applications to gene-discovery and fine-mapping. Based on empirical studies, we show that the proposed gene-based tests are more powerful and help attenuate the confounding effect of LD relative to state-of-the-art gene-based tests. They also have distinct advantages compared with the recently proposed window-based test, Knockoff-Screen, in terms of functional interpretation, and appropriate FDR control at the gene level. Indeed, our simulation results suggest that the knockoff filter procedure needs to be performed at the gene rather than window level if our interest is in identifying genes and controlling FDR at the gene level.

Our gene-based tests can be seen as complementary to the TWAS approach. Like TWAS, they attempt to incorporate the effect of distal regulatory elements into the test. TWAS however is limited to common eQTLs detectable in reference datasets which appear to account for a minority of GWAS signal^17,18^. In contrast, our approach has the ability to assess the effects of coding, noncoding, rare and common variants, including those with no detectable effects on gene expression, and can scan the gene with varied window sizes. Furthermore, the knockoff framework can attenuate the confounding effect of LD, and is able to produce a narrower list of possible causal genes, likely removing some of the false gene discoveries.

In this paper we have focused on using existing external data on gene-enhancer links, and we recognize the limitations of these databases, both in terms of the accuracy of these links and the number of cell-types with available data. Single-cell Hi-C is an emerging technology that could help overcome issues of tissue heterogeneity and expand these maps across many more cell types^69^.

Like all gene-based tests that incorporate genetic variation in distal regulatory elements, our tests are also susceptible to false-positive associations due to, for example, causal variants residing in putative enhancers that may show significant interactions with promoters of multiple genes based on Hi-C data. Identifying the actual causal gene(s) requires follow-up experimental studies, such as CRISPR gene perturbation experiments^70^.

In summary, we propose comprehensive gene-based tests for common and rare variation, both coding and regulatory variation that are more powerful than competitor gene-based tests in the literature. The GeneScan3DKnock approach is implemented in a computationally efficient R package.

## 4 Methods

### 4.1 Overview of existing gene-based association tests

#### Sequence kernel association test (SKAT) and Burden test

We assume that *n* subjects have been sequenced/genotyped in a region of interest (e.g. a gene), with *p* being the number of variants identified in the region. We denote by *y*_*i*_ the phenotype for the *i*-th subject, and **X**_*i*_ = (*X*_*i*1_, …, *X*_*id*_) be *d* covariates (e.g. age, gender, principal components of ancestry, etc.). The vector of *p* genotypes for subject *i* is denoted by **G**_*i*_ = (*G*_*i*1_, …, *G*_*ip*_), where *G*_*i j*_ = 0, 1, 2 represents the number of minor alleles at variant *j* = 1, …, *p*. We are interested in testing for association between the phenotype and the *p* variants, while adjusting for covariates.

For a continuous phenotype, we consider the classical linear model:

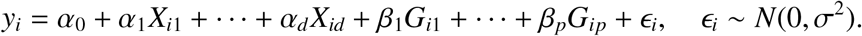

For a binary outcome, the logistic model is considered instead.

The null hypothesis of interest is *H*_0_ : *β*_1_ = … = *β*_*p*_ = 0, i.e. there is no association between the phenotype and any of the *p* variants. To test this hypothesis, the SKAT and Burden test statistics have been proposed^4^. In SKAT, we assume that *β* _*j*_ follow an arbitrary distribution with mean 0 and variance *τw*_*j*_, where *w*_*j*_ is a pre-specified weight for variant *j*; then the null hypothesis *H*_0_ is equivalent to testing *τ* = 0. We conduct a variance-component score test with statistic

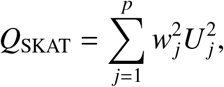

where 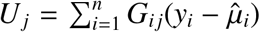 is the score statistic for testing marginal effect of the *j*-th variant (*β* _*j*_ = 0) and 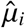 is the fitted value of *y*_*i*_ under the null model (*H*_0_). *Q*_SKAT_ follows a mixture of 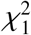 distributions under *H*_0_ and the p-value can be calculated by the Davies method^2,71^. The default way is to assume the weight *w*_*j*_ = Beta(MAF _*j*_; *a*_1_, *a*_2_), where MAF is the minor allele frequency, *a*_1_ = 1 and *a*_2_ = 25; weights based on predicted functional effects of genetic variants are also commonly used^33,72–75^.

A Burden test statistic can also be formulated. If we assume *β* _*j*_ = *w*_*j*_*β*_*c*_, the null hypothesis *H*_0_ above is equivalent to testing *H*_0_ : *β*_*c*_ = 0. The Burden score statistic is defined as:

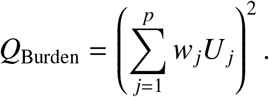

*Q*_Burden_ asymptotically follows a scaled 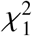 distribution and the p-value can be calculated analytically.

Extensions of these tests to account for sample relatedness and case-control imbalance have also been proposed^6,76^.

#### Transcriptome-wide association tests

A different type of gene-based strategy has become popular in recent years in the context of GWAS studies, namely the transcriptome-wide association studies (TWAS). TWAS integrate large GWAS data and complementary genetic variation-gene expression data (such as GTEx and PsychENCODE). Several TWAS approaches for a single tissue have been proposed, including PrediXcan/MetaXcan and FUSION^14,15,77^. More recently, these approaches have been extended by leveraging the gene expression data across multiple related tissues^78^.

The typical approaches use prediction models, such as Elastic Net or LASSO, to predict the genetically regulated component of gene expression based on genotype information. Specifically, given a gene of interest *k* and *p* cis-SNPs close to the gene, TWAS define the predicted gene expression in a tissue *l* for individual *i* as *Z*_*i,kl*_ = *w*_1,*kl*_*G*_*i*1_ + … + *w*_*p,kl*_*G*_*ip*_ where the weights *w*_*j,kl*_ are the coefficients estimated from genetic variation-gene expression reference datasets such as GTEx. These prediction models are subsequently applied to individual (or summary level) data from GWAS to assess the associations between predicted gene expression and a trait.

### 4.2 Overview of regulatory elements

We describe here the procedure we used to identify proximal and distal regulatory elements for each protein-coding gene.

#### Promoters

Polymerase ChIP-seq data was extracted from the ChIP-Atlas database, a compendium of ChIP-seq data from the NCBI SRA, Roadmap Epigenomics, ENCODE, DNA Data Bank of Japan, and EMBL-EBI ArrayExpress resources, which utilizes MACS2 as the peak identification algorithm^79^. Three sets of peaks were obtained, one using only lung cell lines, one using only neural cell lines, and another set using all cell lines available (tissue classifications for cell lines meeting criteria included Blood, Kidney, Breast, Digestive tract, Uterus, Lung, Neural, Epidermis, Prostate, Pluripotent Stem Cell, Liver, Gonad, Muscle, Adipocyte, Placenta, and Pancreas).

In these polymerase ChIP-seq experiments, the specific antigen targeted corresponded to the ChIP-Atlas polymerase antigen class, which included the following targets: POLR2B, POLR2M, POLR3D, POLR3G, POLR3GL, RNA polymerase II, and RNA polymerase III. Ensembl transcription start sites were extracted using the UCSC ensGene table with the GenomicRanges package genes function, merged against Ensembl IDs, and then again against HUGO gene symbols Bioconductor genome wide annotation for Human (org.Hs.e.g.db). 1 Kb regions upstream of the transcription start site (TSS) that overlapped ChIP-seq peaks were retained for subsequent analysis. Peaks were ranked for each gene according to their proximity to the TSS, the peak narrowness, MACS2 peak score, and the percent overlap with the 1 Kb upstream target peak region. For genes with peaks in the 1 Kb upstream regions, the best peak for each gene was chosen as the highest scoring peak on the five aforementioned metrics. The scoring logic (to choose narrow polymerase peaks that are proximal to and upstream of the TSS) is in line with the size and placement of polymerase peaks in a large scale analysis of mammalian promoters^80^.

#### Enhancers

To define enhancers for a given gene, we use the enhancer elements from GeneHancer (GH) dataset^30^, as well as the activity-by-contact (ABC) model to obtain predicted enhancers for expressed genes^31^. For each gene, we focused on putative enhancers that are outside the gene plus and minus 5 Kb buffer region, and with length less or equal than 10 Kb.

##### GeneHancer Enhancers

To estimate the strength of each gene-enhancer link connection, the gene-enhancer association scores are defined based on five methods: expression quantitative trait loci (eQTLs), enhancer RNA (eRNA) co-expression, transcription factor (TF) coexpression, capture Hi-C (CHi-C) and gene-target distance^30^. Specifically,

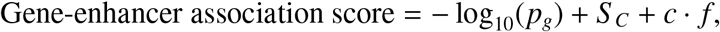

where *p*_*g*_ is the combined p-value for eQTLs, eRNA co-expression and TF co-expression using Fisher’s combination method; *S*_*C*_ is the CHi-C score, constituting the logarithm of the ratio of observed to expected read counts; *c* is a normalizing constant and *f* is the fraction of enhancers in the distance bin out of all genome-wide gene-enhancer pairs^30^.

##### ABC Enhancers

We also incorporate an activity-by-contact (ABC) model to obtain predicted enhancers for expressed genes^31^. The ABC score is constructed for each putative regulatory element E of a gene G as follows:

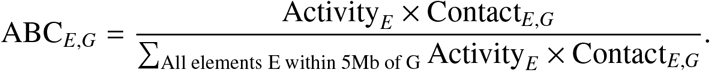

In the ABC model, enhancer activity (Activity_*E*_) is obtained from DNase-seq and H3K27ac ChIP-seq signals, while the Enhancer (E) to Gene (G) contact measure (Contact_*E,G*_) comes from the Knight-Ruiz (KR) normalized, 5 Kb resolution Hi-C state cell types. For each gene, we incorporate the predicted ABC enhancers with ABC scores ≥ 0.02 for 5 cell types and tissues, i.e., K562, GM12878, NCCIT, LNCAP, hepatocytes^31^.

##### Human-brain Hi-C enhancers

For the brain disorder GWAS applications, we leverage Hi-C data for two human brain tissues: adult brain dorsolateral PFC (DLPFC) Hi-C^60^, and Fetal brain (developing cortex) Hi-C with two layers germinal zone (GZ) and cortical and subcortical plate (CP)^61^. The Hi-C data (10Kb resolution) are bias corrected and normalized under iterative correction and eigenvector decomposition (ICE) using hiclib^81^.

We use the Fit-Hi-C method to identify statistically significant chromatin contacts from the given Hi-C matrices. Fit-Hi-C uses spline models to estimate expected contact probabilities using genomic distance and calculated ICE biases. Statistical significance of interactions is calculated using a binomial distribution and p-values corrected for multiple testing^62,63^. We obtain long-range chromatin interactions between 20Kb lower bound and 2Mb upper bound of interaction distances using Fit-Hi-C2 (version 2.0.7)^82^. Hi-C contacts with FDR < 0.01 were selected as significant interactions. Significant Hi-C interacting regions were overlapped with GENCODE v26 promoter coordinates (defined as 2kb upstream and downstream of the TSS) to identify promoter-based interactions.

Additionally, we also incorporate promoter-based interactions as described in^60,61^, by generating background Hi-C interaction profiles from randomly selected regions matched length and GC content to promoters and fitting Hi-C contacts to Weibull distributions. We leverage both Fit-Hi-C method and promoter-anchored chromatin loops provided by^60,61^ together as long-range Hi-C promoter-enhancer interactions. In total, there are 130,782 enhancers for adult brain (AB) Hi-C, 111,143 enhancers for fetal brain Hi-C germinal zone (FB-GZ) and 109,298 enhancers for fetal brain Hi-C cortical and subcortical plate (FB-CP). All putative brain-tissue Hi-C enhancers are in 10Kb resolution.

### 4.3 GeneScan3DKnock: Proposed gene-based association test incorporating regulatory elements and knockoff statistics

We describe here the details of the proposed gene-based test that aims to comprehensively evaluate the effects of rare and common, coding, proximal and distal regulatory variation on a trait of interest. A workflow depicting the overall gene-based testing approach proposed here is shown in Figure 1.

#### Joint testing of rare and common variation within a window

For a fixed window Φ, we incorporate several recent advances for association tests for sequencing studies to compute the corresponding p-value *p*_Φ_, as follows.

For each window we conduct:

a. Burden and SKAT tests for common and low frequency variants 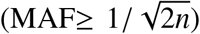 with Beta(MAF _*j*_; 1, 25) weights. These tests aim to detect the combined effect of common and low frequency variants.
b. Burden and SKAT tests for rare variants (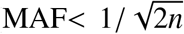 *&* MAC (minor allele count) ≥ *t*) with Beta(MAF _*j*_; 1, 25) weights. These tests aim to detect the combined effect of rare variants.
c. Burden and SKAT tests for rare variants, weighted by cell-type specific functional annotations. These tests aim to utilize functional annotations for improved power^26,83^.
d. Burden test for aggregation of ultra-rare variants (MAC< *t*). These tests aim to aggregate effects from extremely rare variants (e.g. singletons, doubletons, etc.).
e. Single variant score tests for common, low frequency and rare variants (MAC ≥ *t*) in the window.

We then apply the aggregated Cauchy association test^24^ to combine p-values from tests in a-e to compute p-values of each 1D window Φ for all variants, including common and rare variants. Note that for the current analyses we use MAC threshold 10.

#### GeneScan3D: Integrating proximal and distal regulatory elements for a gene

For a given gene *G*, we consider the gene body (i.e. the interval between the transcription start site (TSS) and the end of 3’ UTR) plus and minus 5 Kb buffer regions and integrate a single ChIP-seq promoter and *R* putative enhancers into the analyses (Figure 1). A set of overlapping 1D windows Φ_*m*_, *m* = 1, …, *M* with window sizes 1-5-10 Kb are generated to scan the gene and buffer regions together (each 1D window is overlapping with half of its adjacent windows for a given window size). Then we construct 3D windows for the gene by adding a ChIP-seq promoter and *R* putative enhancers to each 1D window Φ_*m*_, *m* = 1, …, *M* as follows (details on how to identify the regulatory elements for each gene are in Section 4.2):

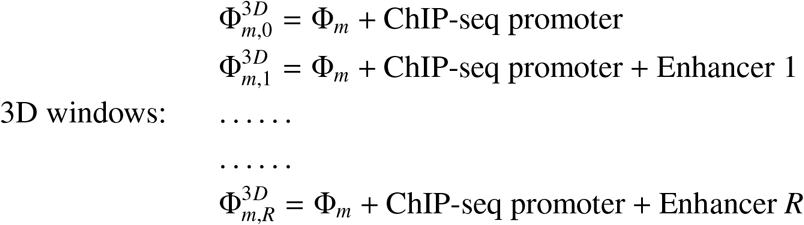

For each such 3D window, we compute a p-value 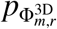, with 1 ≤ *m* ≤ *M*, 0 ≤ *r* ≤ *R* using the proposed combined test for a window. Finally, we compute a gene-level p-value *p*_*G*_ by combining the (1 + *R*) × *M* p-values using the Cauchy’s combination method^24^, as follows:

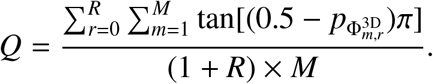

The p-value of the Cauchy statistic is *p*_*G*_ = 1/2 − arctan(*Q*)/π.

#### GeneScan3DKnock: Knockoff-enhanced gene-based test for causal gene discovery

An advantage of the proposed GeneScan3D test is that it allows the discovery of multiple possible causal genes by incorporating information from proximal and distal regulatory elements. However, it is likely that some of those genes are false positives owing to confounding due to linkage disequilibrium (LD) and/or co-regulation. Extensive LD at a locus of interest can confound the results and lead to many genes being significant. For example, if the LD region overlaps several enhancers, all genes regulated by such enhancers may show a significant signal. The knockoff framework^28^, a recent advance in statistics, can be leveraged to reduce the effect of LD in such cases, and can help prioritize a narrow list of potential causal genes. Furthermore, the knockoff-based test is of independent interest, as by design it controls the FDR at a target level under arbitrary correlation structure, and can have higher power to identify additional significant genes that are missed by the conventional gene-based test as we show empirically in the applications. The knockoff-based test has two steps: the knockoff generation, and the filtering of the results using the knockoff filter.

##### Model-X Knockoff generation

The idea of the knockoff-based procedure is to generate artificial or knockoff genotypes 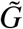 such that for any subset *K* of variants the distribution of 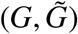 is invariant when swapping *G*_*K*_ and 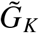, i.e. 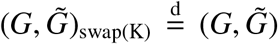. Additionally the knockoff genotypes have the property that 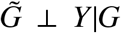. Note that the well-known permutation procedure that permutes the samples does not guarantee these exchangeability properties between the original and knockoff genotypes. To generate valid knockoff genotypes we can use a sequential model for knockoff generation that leverages the local patterns of linkage disequilibrium, as previously proposed based on the Hidden Markov Models (HMMs)^32,84^ or an auto-regressive model^8^, in such a way that the knockoff genotypes are exchangeable with the original (true) genotypes *G* but are independent of the phenotype conditional on the original genotypes. The knockoff genotypes serve as negative controls and are designed to mimic the correlation or LD structure found within the original genotypes. Specifically, we sequentially sample for each variant *j* the corresponding knockoff genotype 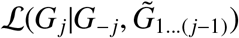, independent of the observed value of *G*_*j*_. Because of the HMM’s significant computational complexity with unphased genetic data, in order to generate knockoff genotypes we rely on a recently introduced, computationally efficient auto-regressive model that follows from the assumption that genotypes can be approximately modeled by a multivariate normal distribution:

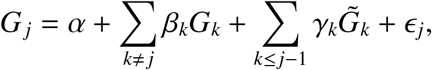

where ϵ _*j*_ is a random error term. (Note that we can leverage the approximate block structure for LD in the genome to only include variants in a neighborhood of the current variant *j*.) We estimate (*α, β, γ*) by minimizing the mean squared loss. We calculate the residual 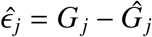 and its permutation 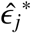, and then we define the knockoff feature as 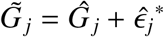. More details on this knockoff generation procedure, and its theoretical and empirical properties can be found in^8^.

##### Knockoff filter

Once the knockoff genotypes 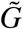 are generated, the knockoff filter is used to select significant genes. Specifically, we perform a gene-based test as described above (GeneScan3D) in both the original cohort and the knockoff one. Let *p*_*G*_, and 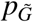 be the resulting p-values. We define a feature statistic by contrasting the observed p-value for each gene to its counterpart based on the knockoff data. More precisely, the feature statistic for a gene *G* is defined as 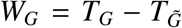, where *T*_*G*_ = − log_10_(*p*_*G*_) and 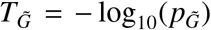 are the importance score for gene *G* in the original and knockoff cohort, respectively. This feature statistic has the flip-sign property, meaning that swapping the genetic variants in gene *G* with their knockoff counterparts changes the sign of *W*_*G*_. A data-adaptive threshold *τ* for *W*_*G*_ can be determined by the knockoff filter^28^ so that the FDR is controlled at the nominal level *q*, as follows:

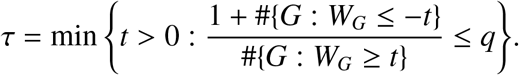

We select all genes with *W*_*G*_ ≥ *τ* since genes with large feature statistics are more likely to be causal (non-null) genes. This follows from the exchangeability property between the original and the knockoff genotypes, which ensures that the importance scores (*T*_*G*_ and 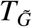) for the null genes are exchangeable, and therefore the feature statistic *W*_*G*_ is symmetric around 0 for the null genes, but tends to be larger than 0 for non-null genes.

We additionally compute the corresponding q-value for a gene, *q*_*G*_. The q-value already incorporates correction for multiple testing, and is defined as the minimum FDR that can be attained when all tests showing evidence against the null hypothesis at least as strong as the current one are declared as significant. In particular, we define the q-value for a gene *G* with feature statistic *W*_*G*_ > 0 as

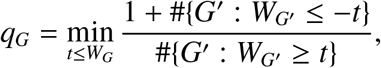

where 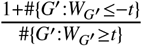 is an estimate of the proportion of false discoveries if we were to select all genes with feature statistic > *t* (with *t* > 0). For genes with feature statistic *W*_*G*_ ≤ 0 we set q_*G*_ =1.

##### Multiple knockoffs

To improve the power and stability of the knockoff procedure we implement a multiple knockoff procedure^8,85^ where the inference is based on generating multiple, independent knockoff datasets. Gimenez and Zou^85^ proposed an extension of the sequential model for knockoff generation to multiple knockoffs and showed the validity of the multiple knockoff generation procedure in controlling the FDR. We implement this procedure here to generate multiple independent knockoff datasets. Briefly, we sequentially sample for each variant 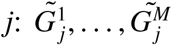 from 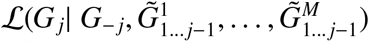, where *M* is the number of knockoffs. With multiple knockoffs, the feature statistic for a gene *G* is defined as

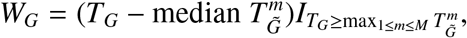

where 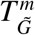 is the gene importance score for gene *G* in the *m*-th knockoff replicate, and *I* is an indicator function. We define

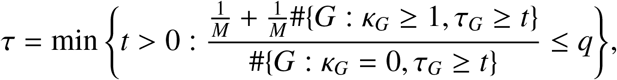

where 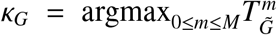 (note that 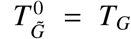) and 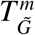. We select genes with *W*_*G*_ ≥ *τ*, i.e. those genes that have importance scores greater than any of those corresponding to the *M* knockoffs (*κ*_*G*_ = 0), and for which the difference from the median importance score is above some threshold (*τ*_*G*_ ≥ *τ*). A q-value for a gene *G* can be computed for the multiple knockoff scenario, similar to the single knockoff case.

The multiple knockoff procedure helps improve power because at a target FDR of *q*, the single knockoff approach needs to make a minimum of 1/*q* discoveries while a multiple knockoff approach with *M* knockoffs decreases this detection threshold to 1/*Mq*. Therefore, in situations where the signal is sparse and the target FDR level *q* is low, a single knockoff procedure will have very low power. In such cases, the multiple knockoff procedure will tend to improve power. Furthermore, the multiple knockoff procedure also helps with improving the stability of the selected genes given that each knockoff generation is random, and therefore the results from a single knockoff can be unstable.

#### Additional tests for comparison

We compare with the nearest competitor gene-based tests in the literature (MAGMA/H-MAGMA, STAAR-O) as well as with the GeneScan1D test, as described below. We also compare with a recently proposed window-based test, KnockoffScreen.

##### GeneScan1D analysis

We scan the gene *G* and the two 5 Kb buffer regions on either side of the gene using 1D windows with sizes 1-5-10 Kb. For each 1D window, we conduct the combined tests and finally combine p-values for M 1D windows using the Cauchy combination method.

##### MAGMA/H-MAGMA

MAGMA^11^ is a commonly-used gene-based test for GWAS data that links SNPs to their closest gene (e.g. 35 Kb upstream and 10 Kb downstream of each gene). Because of the large number of SNPs and their collinearity, MAGMA is based on a principal component regression model, using the F-test to compute the gene-based p-value. In addition to common variants, it can also incorporate rare variants by computing a burden score for all rare variants linked to a specific gene. Each rare variant is weighted according to 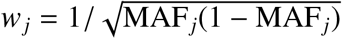. Then MAGMA regresses the dependent variable on the principal components (accounting for 99.9%) of the genetic matrix.

H-MAGMA^12^ is a recent extension of MAGMA that assigns SNPs to genes by using longrange chromatin interactions in disease relevant tissues. Since the current implementation^12^ is focused on human brain tissue, for a fair comparison, in the analyses presented in this paper we use the same method to assign SNPs in putative regulatory elements to their cognate genes as employed for the GeneScan3D test.

##### STAAR-O

STAAR-O^13^ is a recently proposed rare variant (RV) association test, incorporating functional annotations and genetic variation in distal regulatory elements. The STAAR-O framework first calculates a set of test statistics including STAAR-Burden, STAAR-SKAT and STAAR-ACAT-V using different annotation weights. The aggregated Cauchy association test (ACAT) method is used to combine the resulting P-values into the STAAR-Omnibus test statistic. In addition to a sliding window approach, STAAR-O can also perform gene-centric analyses by generating p-values for the gene body and associated regulatory elements.

Although STAAR-O focuses primarily on detecting rare variant associations, for a fair comparison with our proposed test we apply STAAR-O to all variants, and only common variants using the same MAF/MAC thresholds as used for our proposed tests (MAF threshold 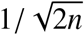 and MAC threshold 10). Similarly, for STAAR-SKAT and STAAR-Burden tests, we use the same weighting scheme for the variants (Beta(MAF _*j*_; 1, 25)), and for STAAR-ACAT-V that aggregates p-values from the extremely rare variants of Burden test and the individual variant score tests, we use weights Beta(MAF;1,1), as employed in GeneScan3D. We also use the same regulatory elements and functional annotations that we used for the GeneScan3D test. For each protein-coding gene, three gene functional categories are considered: (1) The entire gene body (2) ChIP-seq promoter; and (3) *R* putative GH and ABC enhancers together. Finally, we combine the STAAR-O p-values for the 3 categories using the Cauchy combination method.

For the sake of completeness, we also apply the default setting of STAAR-O (R package v.0.9.5) using both weights Beta(MAF _*j*_; 1, 25) and Beta(MAF _*j*_; 1, 1); MAF threshold 0.05 for simulations and MAF threshold 0.01 for real data analysis; MAC threshold 10 for STAAR-ACAT-V, for RVs association studies^13^. We note however that when causal variants include common variants as in our settings, this default setting may be less powerful.

##### KnockoffScreen

Although our main comparisons are with gene-based tests, we perform additional comparisons with the recently proposed window-based method KnockoffScreen^8^ in order to illustrate the need for a gene-based knockoff filter when our interest is controlling the FDR at the gene level. For KnockoffScreen, we scan each locus using several window sizes, with half of each window overlapping the adjacent windows of the same size. For each window, we conduct joint testing of rare and common variation using original genotype and *M* = 1, 3, 5 knockoffs. Then we use the knockoff filter to detect windows that are significant at a specified FDR target level. Being a window-based method, KnockoffScreen is designed to identify significant windows at a specified target level. Because here genes are the functional units of interest, we estimate the empirical power and FDR in terms of genes. Specifically, we define a gene as detected by KnockoffScreen if there is at least one detected window that overlaps the gene and/or its regulatory elements.We compute the gene-level empirical power as the proportion of causal genes being detected and the empirical FDR as the proportion of the detected genes that are noise. Note that the empirical gene-level FDR does not correspond to the window-level FDR that KnockoffScreen is aiming to control at a target level. The empirical window-level FDR is computed as the proportion of detected windows not overlapping with the causal loci and the empirical power is defined as the proportion of causal loci being identified (i.e. the causal locus overlaps with at least one detected window).

### 4.4 Software implementation

We have developed a computationally efficient R package, GeneScan3DKnock (https://github.com/Iuliana-Ionita-Laza/GeneScan3DKnock), to facilitate the gene-based test analyses described here. The package implements the 1D and 3D versions of the gene-based tests, along with the knockoff-enhanced tests. The computational time of GeneScan3DKnock depends on the sample size, gene size and the number of regulatory elements. To perform WGS gene-based tests on ∼ 4000 individuals using the ADSP data required 27h for 22 2.60 GHz computing cores with 70 gigabyte memory for AD trait. The TOPMed COPDgene projects with ∼ 5600 Non Hispanic White individuals required 30h for 22 2.50 GHz computing cores with 70 gigabyte memory for each trait.

#### Visualization of the promoter-enhancer link data for the significant genes in the 3D analyses

It is also of interest to visualize the promoter-enhancer links for genes with significant 3D test results. For the GeneScan3D analysis, each gene is comprised of *M* × (1 + *R*) 3D windows, including *M* 1D windows + promoter, and *M* 1D windows + promoter + one of *R* enhancers (Section 4.3). For each significant gene in the GeneScan3D test, we can visualize the promoter-enhancer links corresponding to the 3D window with minimum p-value using the Gviz and GenomicInteractions packages in R. The code to generate the plots is available as a vignette (https://github.com/Iuliana-Ionita-Laza/GS3DKViz).

## Data Availability

We used data from existing studies from COPDGene (TopMED, dbGaP phs000951.v4.p4) and the Alzheimer’s Disease Sequencing Project (dbGaP phs000572.v8.p4), and summary level GWAS results on neuropsychiatric and neurodegenerative traits are available from^51–59^.

## Data Availability

The data used are available on dbGAP.

## Code Availability

We have implemented GeneScan3DKnock in a computationally efficient R package that can be applied generally to the analysis of other whole-genome sequencing or GWAS studies.

## Acknowledgements

This research was supported by NIH/NIMH awards MH106910 and MH095797 (IIL) and NIH/NIA award AG066206 (ZH). We gratefully acknowledge the studies and participants who provided biological samples and data for the ADSP and TOPMed projects. The full study-specific acknowledgements are detailed in the Supplementary Note.

## Web-based resources

ABC enhancers: https://www.engreitzlab.org/resources/

BioBank Japan: http://jenger.riken.jp/en/result

BioBank Japan Browser (GeneScan3D): http://www.funlda.com/gs3d/bbj

ChIP-Atlas: https://chip-atlas.org/

GeneHancer enhancers: http://www.genecards.org/

GeneScan3DKnock: https://github.com/Iuliana-Ionita-Laza/GeneScan3DKnock

GS3DKViz: https://github.com/Iuliana-Ionita-Laza/GS3DKViz

GenomicInteractions: https://www.rdocumentation.org/packages/GenomicInteractions

GenoNet: http://www.funlda.com/genonet

GTEx portal: https://gtexportal.org/home/

Gviz: https://www.rdocumentation.org/packages/Gviz

MetaXcan: https://github.com/hakyimlab/MetaXcan

Pan-UK Biobank Summary Statistics: https://pan.ukbb.broadinstitute.org/

TOPMed freeze5b dataset: https://www.nhlbiwgs.org/topmed-whole-genome-sequencing-project-freeze-5b-phases-1-and-2

UCSC genome browser: https://genome.ucsc.edu/

UK Biobank Summary Statistics: ftp://share.sph.umich.edu/UKBB

SAIGE HRC/ UK Biobank Results (GeneScan3D): https://doi.org/10.5281/zenodo.4477703

UK Biobank Browser (GeneScan3D): http://www.funlda.com/gs3d

## Supplemental Material

### Whole-genome sequencing data and analyses

#### The Alzheimer’s Disease Sequencing Project (ADSP)

The ADSP data include 3,085 whole genomes from the ADSP Discovery Extension Study including 1,096 Non-Hispanic White (NHW), 977 African American (AA) descent and 1,012 Caribbean Hispanic (CH). Sequencing for these samples was conducted through three National Human Genome Research Institute (NHGRI) funded Large Scale Sequencing and Analysis Centers (LSACs): Baylor College of Medicine Human Genome Sequencing Center, the Broad Institute, the McDonnell Genome Institute at Washington University. The samples were sequenced on the Illumina HiSeq X Ten platform with 150bp paired-end reads. Additionally, the dataset includes 809 whole genomes from the Alzheimer’s Disease Neuroimaging Initiative (ADNI) with 756 NHW, 28 AA and 25 others. The samples were sequenced on the Illumina HiSeq 2000 platform with 100bp paired-end reads. Whole-genome sequence data on 809 ADNI subjects (cases, mild cognitive impairment, and controls) have been harmonized using the ADSP pipeline for joint analysis. The ADSP Quality Control Work Group performs QC and concordance checks into an overall ADSP VCF file.

#### COPDGene from the TOPMed Project

Eligible subjects in COPDGene Study (NCT00608764, www.copdgene.org) were of non-Hispanic white (NHW) or African-American (AA) ancestry, aged 45-80 years old, with at least 10 pack-years of smoking and no diagnosed lung disease other than COPD or asthma. IRB approval was obtained at all study centers, and all study participants provided written informed consent. All subjects underwent a baseline survey, including demographics, smoking history, and symptoms; preand post-bronchodilator lung function testing; and chest CT scans. Samples from COPDGene were sequenced at the Broad Institute and at the Northwest Genomics Center at the University of Washington. Variants for all TOPMed samples were jointly called by the Informatics Research Center at the University of Michigan. For details on sequencing and variant calling methods, see https://www.nhlbiwgs.org/topmed-whole-genome-sequencing-project-freeze-5b-phases-1-and-2. QC included comparison of annotated and genetic sex and comparison of genotypes from prior SNP array data with genotypes called from sequencing. Samples with questionable identity from either of these checks were excluded from analysis.

#### Acknowledgements

The Alzheimer’s Disease Sequencing Project (ADSP) is comprised of two Alzheimer’s Disease (AD) genetics consortia and three National Human Genome Research Institute (NHGRI) funded Large Scale Sequencing and Analysis Centers (LSAC). The two AD genetics consortia are the Alzheimer’s Disease Genetics Consortium (ADGC) funded by NIA (U01 AG032984), and the Cohorts for Heart and Aging Research in Genomic Epidemiology (CHARGE) funded by NIA (R01 AG033193), the National Heart, Lung, and Blood Institute (NHLBI), other National Institute of Health (NIH) institutes and other foreign governmental and non-governmental organizations. The Discovery Phase analysis of sequence data is supported through UF1AG047133 (to Drs. Schellenberg, Farrer, Pericak-Vance, Mayeux, and Haines); U01AG049505 to Dr. Seshadri; U01AG049506 to Dr. Boerwinkle; U01AG049507 to Dr. Wijsman; and U01AG049508 to Dr. Goate and the Discovery Extension Phase analysis is supported through U01AG052411 to Dr. Goate, U01AG052410 to Dr. Pericak-Vance and U01 AG052409 to Drs. Seshadri and Fornage. Data generation and harmonization in the Follow-up Phases is supported by U54AG052427 (to Drs. Schellenberg and Wang). The ADGC cohorts include: Adult Changes in Thought (ACT), the Alzheimer’s Disease Centers (ADC), the Chicago Health and Aging Project (CHAP), the Memory and Aging Project (MAP), Mayo Clinic (MAYO), Mayo Parkinson’s Disease controls, University of Miami, the Multi-Institutional Research in Alzheimer’s Genetic Epidemiology Study (MIRAGE), the National Cell Repository for Alzheimer’s Disease (NCRAD), the National Institute on Aging Late Onset Alzheimer’s Disease Family Study (NIA-LOAD), the Religious Orders Study (ROS), the Texas Alzheimer’s Research and Care Consortium (TARC), Vanderbilt University/Case Western Reserve University (VAN/CWRU), the Washington Heights-Inwood Columbia Aging Project (WHICAP) and the Washington University Sequencing Project (WUSP), the Columbia University Hispanic- Estudio Familiar de Influencia Genetica de Alzheimer (EFIGA), the University of Toronto (UT), and Genetic Differences (GD).

The CHARGE cohorts are supported in part by National Heart, Lung, and Blood Institute (NHLBI) infrastructure grant HL105756 (Psaty), RC2HL102419 (Boerwinkle) and the neurology working group is supported by the National Institute on Aging (NIA) R01 grant AG033193. The CHARGE cohorts participating in the ADSP include the following: Austrian Stroke Prevention Study (ASPS), ASPS-Family study, and the Prospective Dementia Registry-Austria (ASPS/PRODEM-Aus), the Atherosclerosis Risk in Communities (ARIC) Study, the Cardiovascular Health Study (CHS), the Erasmus Rucphen Family Study (ERF), the Framingham Heart Study (FHS), and the Rotterdam Study (RS). ASPS is funded by the Austrian Science Fond (FWF) grant number P20545-P05 and P13180 and the Medical University of Graz. The ASPS-Fam is funded by the Austrian Science Fund (FWF) project I904),the EU Joint Programme -Neurodegenerative Disease Research (JPND) in frame of the BRIDGET project (Austria, Ministry of Science) and the Medical University of Graz and the Steiermrkische Krankenanstalten Gesellschaft. PRODEM-Austria is supported by the Austrian Research Promotion agency (FFG) (Project No. 827462) and by the Austrian National Bank (Anniversary Fund, project 15435. ARIC research is carried out as a collaborative study supported by NHLBI contracts (HHSN268201100005C, HHSN268201100006C, HHSN268201100007C, HHSN268201100008C, HHSN268201100009C, HHSN268201100010C, HHSN268201100011C, and HHSN268201100012C). Neurocognitive data in ARIC is collected by U01 2U01HL096812, 2U01HL096814, 2U01HL096899, 2U01HL096902, 2U01HL096917 from the NIH (NHLBI, NINDS, NIA and NIDCD), and with previous brain MRI examinations funded by R01-HL70825 from the NHLBI. CHS research was supported by contracts HHSN268201200036C, HHSN268200800007C, N01HC55222, N01HC85079, N01HC85080, N01HC85081, N01HC85082, N01HC85083, N01HC85086, and grants U01HL080295 and U01HL130114 from the NHLBI with additional contribution from the National Institute of Neurological Disorders and Stroke (NINDS). Additional support was provided by R01AG023629, R01AG15928, and R01AG20098 from the NIA. FHS research is supported by NHLBI contracts N01-HC-25195 and HHSN268201500001I. This study was also supported by additional grants from the NIA (R01s AG054076, AG049607 and AG033040 and NINDS (R01 NS017950). The ERF study as a part of EUROSPAN (European Special Populations Research Network) was supported by European Commission FP6 STRP grant number 018947 (LSHG-CT-2006-01947) and also received funding from the European Community’s Seventh Framework Programme (FP7/2007-2013)/grant agreement HEALTH-F4-2007-201413 by the European Commission under the programme “Quality of Life and Management of the Living Resources” of 5th Framework Programme (no. QLG2-CT-2002-01254). High-throughput analysis of the ERF data was supported by a joint grant from the Netherlands Organization for Scientific Research and the Russian Foundation for Basic Research (NWO-RFBR 047.017.043). The Rotterdam Study is funded by Erasmus Medical Center and Erasmus University, Rotterdam, the Netherlands Organization for Health Research and Development (ZonMw), the Research Institute for Diseases in the Elderly (RIDE), the Ministry of Education, Culture and Science, the Ministry for Health, Welfare and Sports, the European Commission (DG XII), and the municipality of Rotterdam. Genetic data sets are also supported by the Netherlands Organization of Scientific Research NWO Investments (175.010.2005.011, 911-03-012), the Genetic Laboratory of the Department of Internal Medicine, Erasmus MC, the Research Institute for Diseases in the Elderly (014-93-015; RIDE2), and the Netherlands Genomics Initiative (NGI)/Netherlands Organization for Scientific Research (NWO) Netherlands Consortium for Healthy Aging (NCHA), project 050-060-810. All studies are grateful to their participants, faculty and staff. The content of these manuscripts is solely the responsibility of the authors and does not necessarily represent the official views of the National Institutes of Health or the U.S. Department of Health and Human Services. The four LSACs are: the Human Genome Sequencing Center at the Baylor College of Medicine (U54 HG003273), the Broad Institute Genome Center (U54HG003067), The American Genome Center at the Uniformed Services University of the Health Sciences (U01AG057659), and the Washington University Genome Institute (U54HG003079). Biological samples and associated phenotypic data used in primary data analyses were stored at Study Investigators institutions, and at the National Cell Repository for Alzheimer’s Disease (NCRAD, U24AG021886) at Indiana University funded by NIA. Associated Phenotypic Data used in primary and secondary data analyses were provided by Study Investigators, the NIA funded Alzheimer’s Disease Centers (ADCs), and the National Alzheimer’s Coordinating Center (NACC, U01AG016976) and the National Institute on Aging Genetics of Alzheimer’s Disease Data Storage Site (NIAGADS, U24AG041689) at the University of Pennsylvania, funded by NIA, and at the Database for Genotypes and Phenotypes (dbGaP) funded by NIH. This research was supported in part by the Intramural Research Program of the National Institutes of health, National Library of Medicine. Contributors to the Genetic Analysis Data included Study Investigators on projects that were individually funded by NIA, and other NIH institutes, and by private U.S. organizations, or foreign governmental or nongovernmental organizations.

The COPDGene project was supported by Award Number U01 HL089897 and Award Number U01 HL089856 from the National Heart, Lung, and Blood Institute. The content is solely the responsibility of the authors and does not necessarily represent the official views of the National Heart, Lung, and Blood Institute or the National Institutes of Health. COPDGene is also supported by the COPD Foundation through contributions made to an Industry Advisory Board comprised of AstraZeneca, Boehringer-Ingelheim, Genentech, GlaxoSmithKline, Novartis, Pfizer, Siemens, and Sunovion. Molecular data for the Trans-Omics in Precision Medicine (TOPMed) program was supported by the National Heart, Lung and Blood Institute (NHLBI). Genome Sequencing for NHLBI TOPMed: Genetic Epidemiology of COPD (COPDGene) in the TOPMed Program. (phs000951) was performed at the University of Washington Northwest Genomics Center (3R01HL089856-08S1) and the Broad Institute of MIT and Harvard (HHSN268201500014C). Core support including centralized genomic read mapping and genotype calling, along with variant quality metrics and filtering were provided by the TOPMed Informatics Research Center (3R01HL-117626-02S1; contract HHSN268201800002I). Core support including phenotype harmonization, data management, sample-identity QC, and general program coordination were provided by the TOPMed Data Coordinating Center (R01HL-120393; U01HL-120393; contract HHSN268201800001I). We gratefully acknowledge the studies and participants who provided biological samples and data for TOPMed.

**Figure S1:**
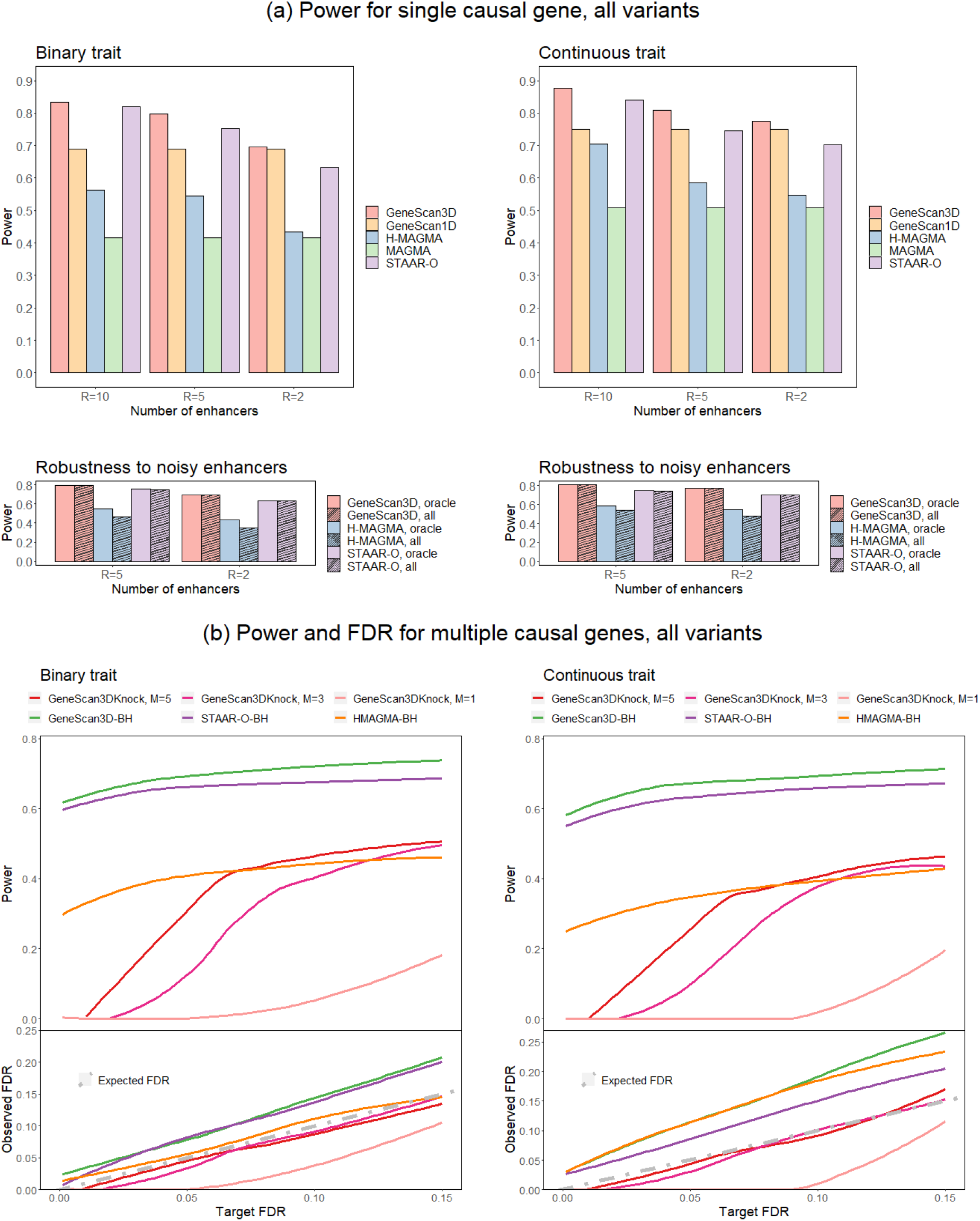
Power and false discovery rate of the proposed gene-based tests, binary and continuous traits with tests including all variants. (a) Power and robustness to noisy enhancers. The top panels show power for the GeneScan3D, GeneScan1D, H-MAGMA, MAGMA and STAAR-O tests. The number of enhancers (*R*) ranges from 2 to 10. The bottom panels show power for the GeneScan3D, H-MAGMA and STAAR-O tests assuming causal variants in *R* = {2, 5} ‘causal’ enhancers. Power is compared between using only the *R* = {2, 5} ‘causal’ enhancers (the oracle approach) vs. using all 10 enhancers (including noisy enhancers). (b) Power and false discovery rate (FDR) for GeneScan3DKnock using different number of knockoffs and the Benjamini-Hochberg (BH) procedure for GeneScan3D, STAAR-O and H-MAGMA.

**Figure S2:**
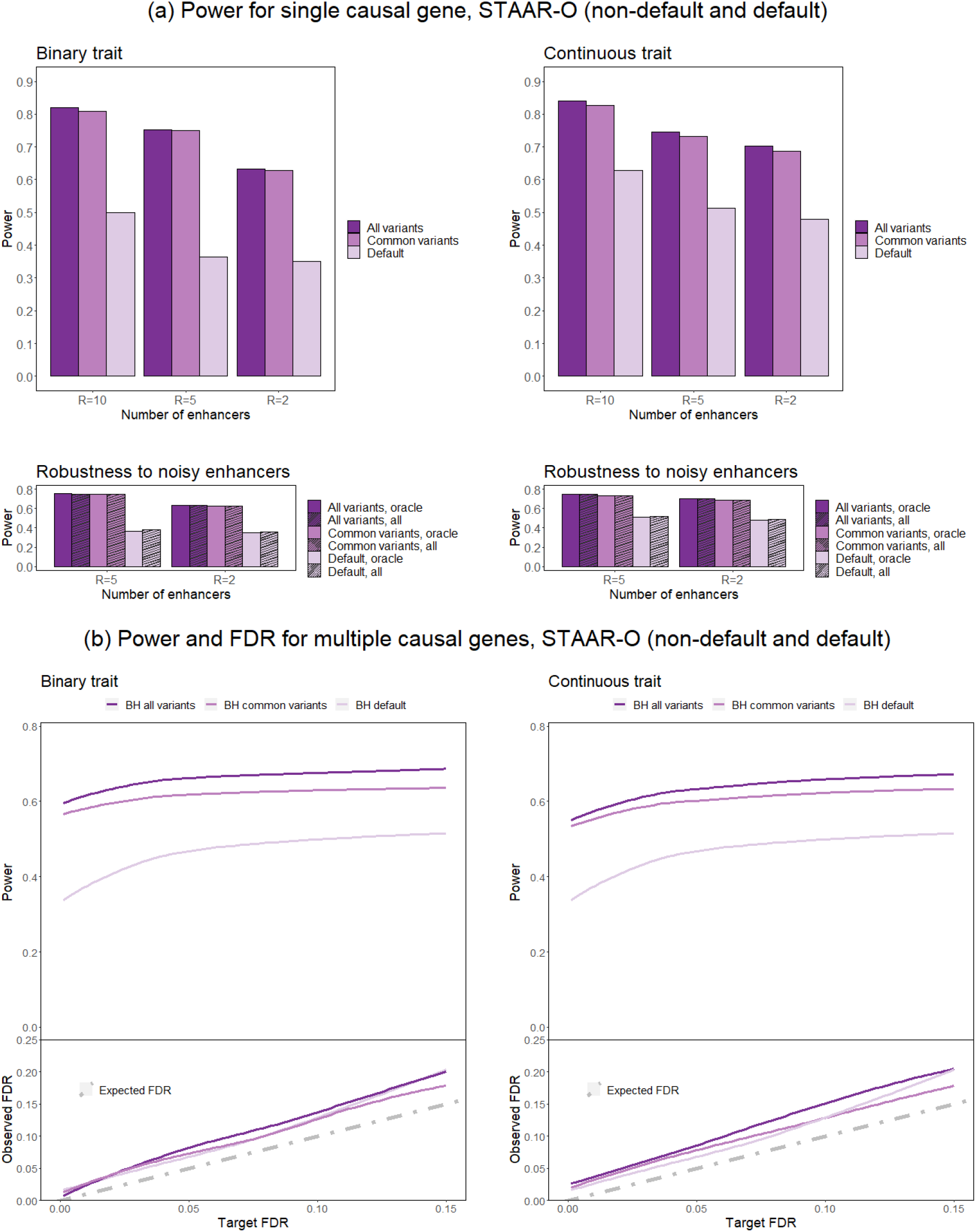
Power and false discovery rate of STAAR-O tests with non-default (all variants and common variants only) and default (rare variants only) settings, binary and continuous traits. (a) Power and robustness to noisy enhancers. The top panels show power for the STAAR-O tests. The number of enhancers (*R*) ranges from 2 to 10. The bottom panels show power for the STAAR-O tests assuming causal variants in *R* = {2, 5} ‘causal’ enhancers. Power is compared between using only the *R* = {2, 5} ‘causal’ enhancers (the oracle approach) vs. using all 10 enhancers (including noisy enhancers). (b) Power and false discovery rate (FDR) for the Benjamini-Hochberg (BH) procedure for STAAR-O.

**Figure S3:**
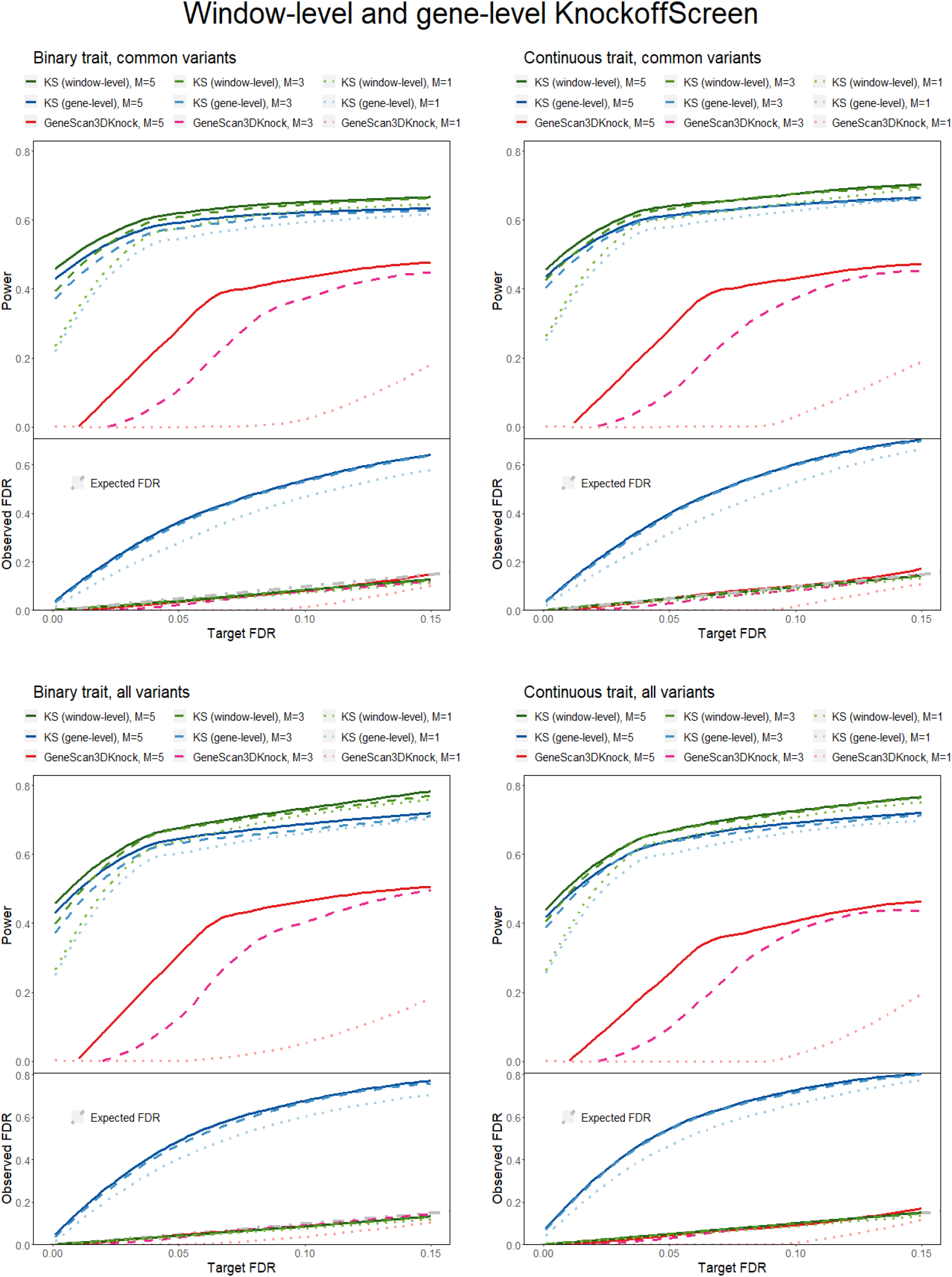
Power and false discovery rate of window-level KnockoffScreen, gene-level KnockoffScreen, and GeneScan3DKnock, binary and continuous traits with tests including common variants only and all variants. Power and FDR are reported when using different number of knockoffs.

**Figure S4:**
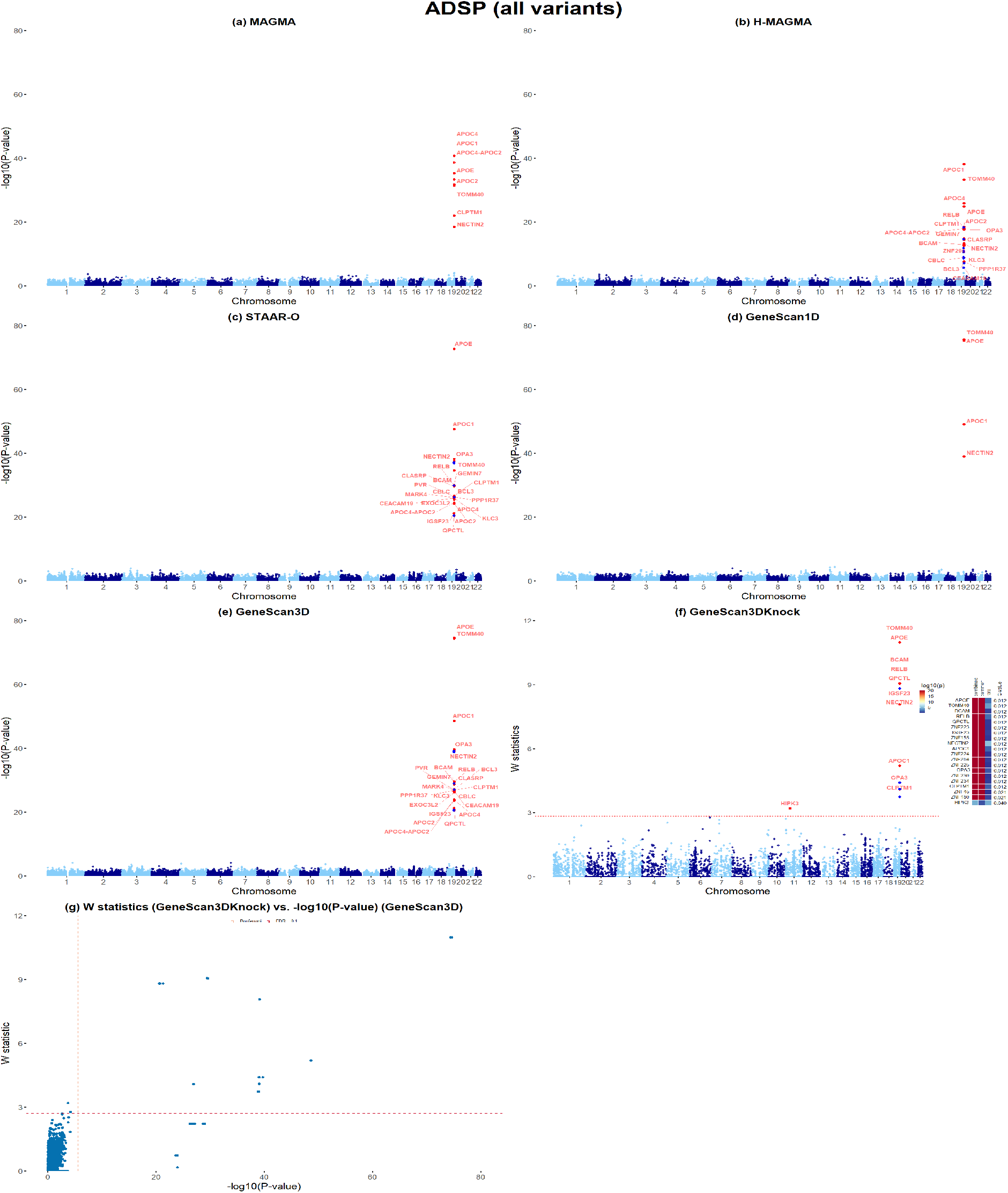
Applications to ADSP whole-genome sequencing data, all variants. (a)-(e) Manhattan plots of MAGMA, H-MAGMA, STAAR-O, GeneScan1D and GeneScan3D results, respectively. (f) Manhattan plot of GeneScan3DKnock results. Genes within the zinc-finger-containing (ZNF) gene cluster on chr19 are unlabeled and shown in blue in H-MAGMA, GeneScan3D and GeneScan3DKnock analyses for clear visualization. The right panel shows a heatmap with p-values of GeneScan3D test (truncated at 10^−20^) for all genes passing the FDR=0.1 threshold, and the corresponding q-values that already incorporate correction for multiple testing. The genes are shown in descending order of the knockoff statistics. (g) Scatter plot of *W* knockoff statistics (GeneScan3DKnock) vs. − log_10_(p value) (GeneScan3D) for all variants. Each dot represents a gene. The dashed lines show the significance threshold defined by Bonferroni correction (for p-values), and the data-adaptive threshold for false discovery rate control (FDR; for W statistic).

**Figure S5:**
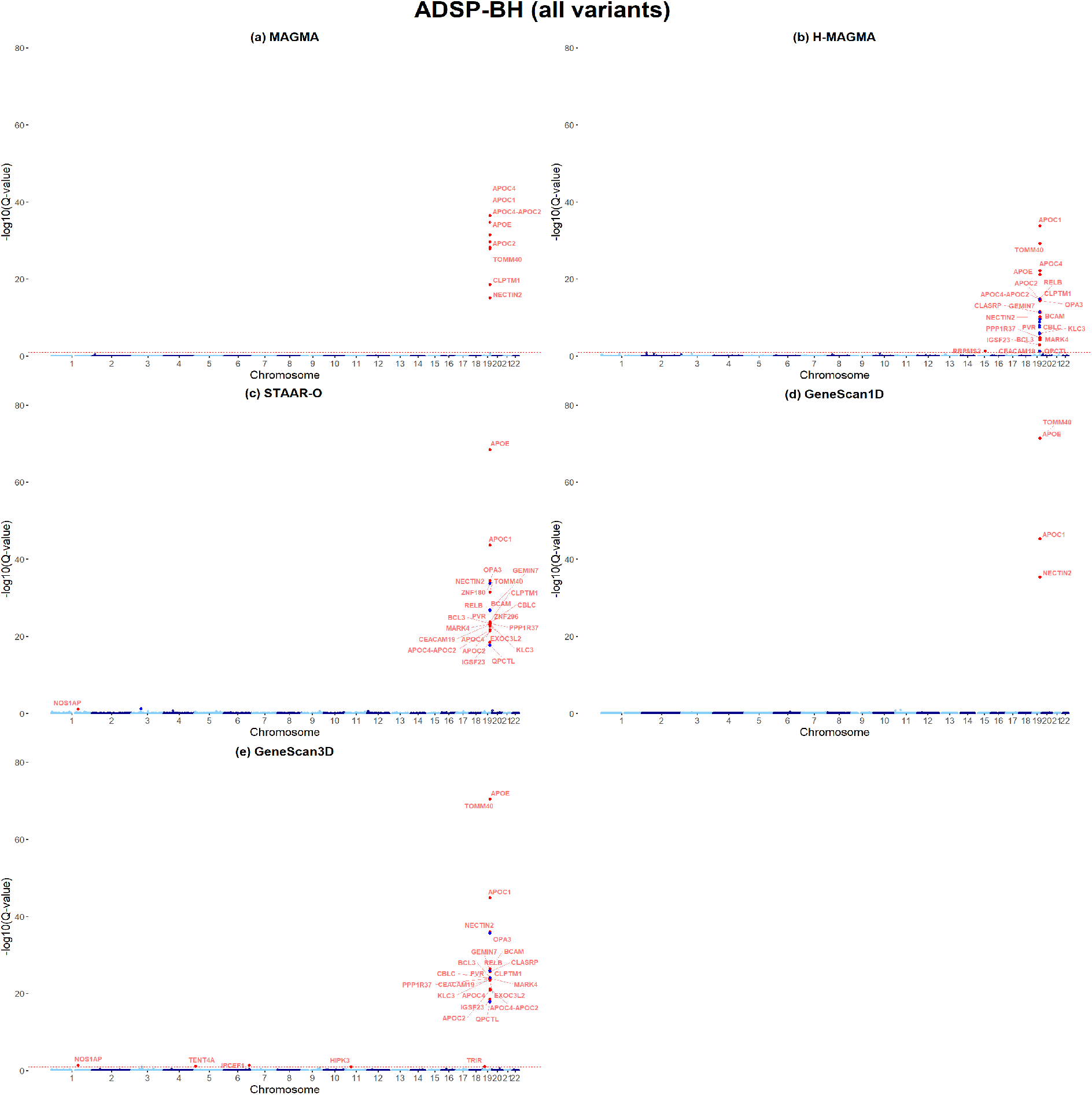
Applications to ADSP whole-genome sequencing data, all variants. (a)-(e) Manhattan plots of MAGMA, H-MAGMA, STAAR-O, GeneScan1D and GeneScan3D and GeneS-can3DKnock results using BH for FDR control, respectively.

**Figure S6:**
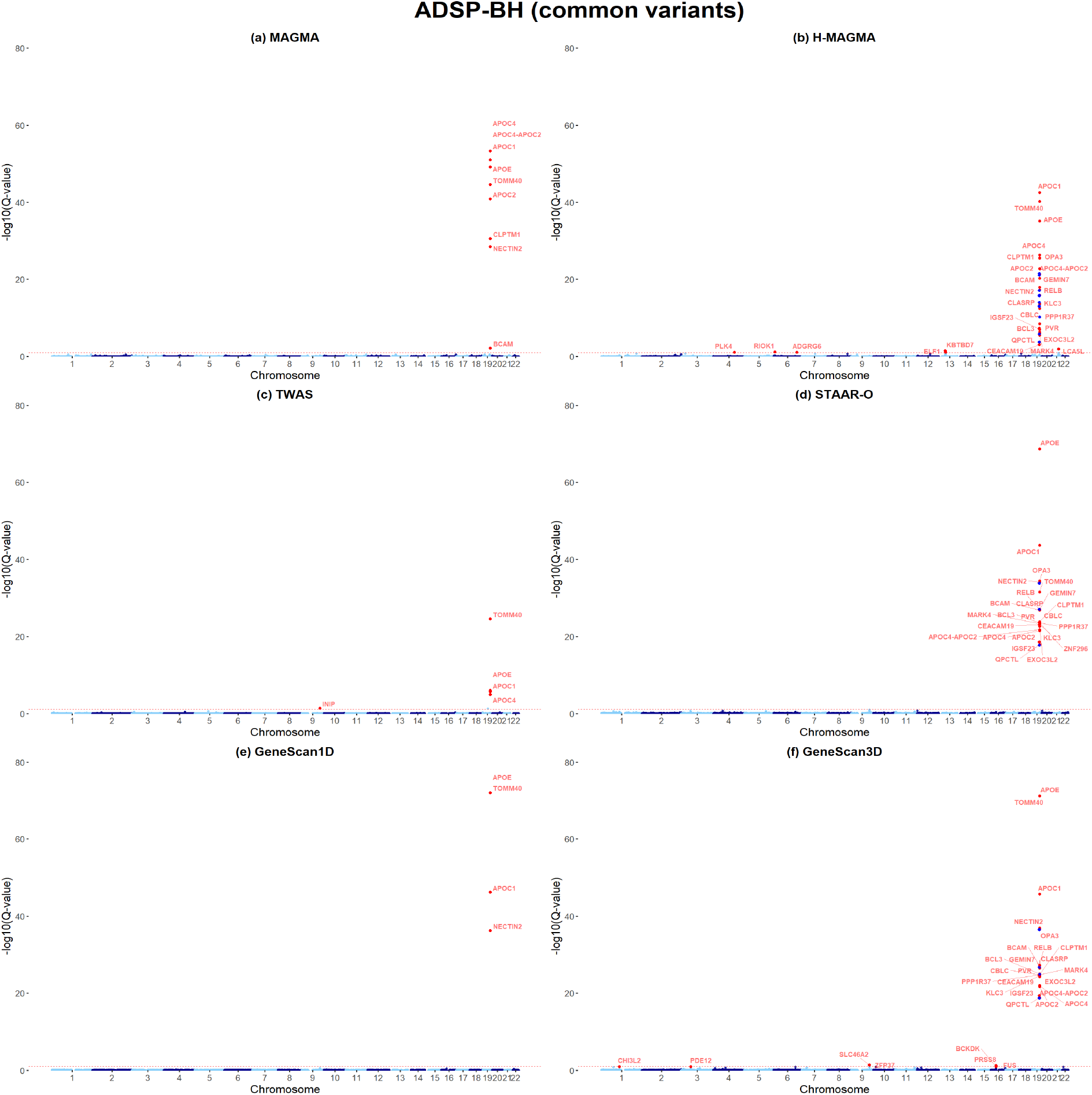
Applications to ADSP whole-genome sequencing data, common variants. (a)-(f) Manhattan plots of MAGMA, H-MAGMA, TWAS, STAAR-O, GeneScan1D and GeneScan3D and GeneScan3DKnock results using BH for FDR control, respectively.

**Figure S7:**
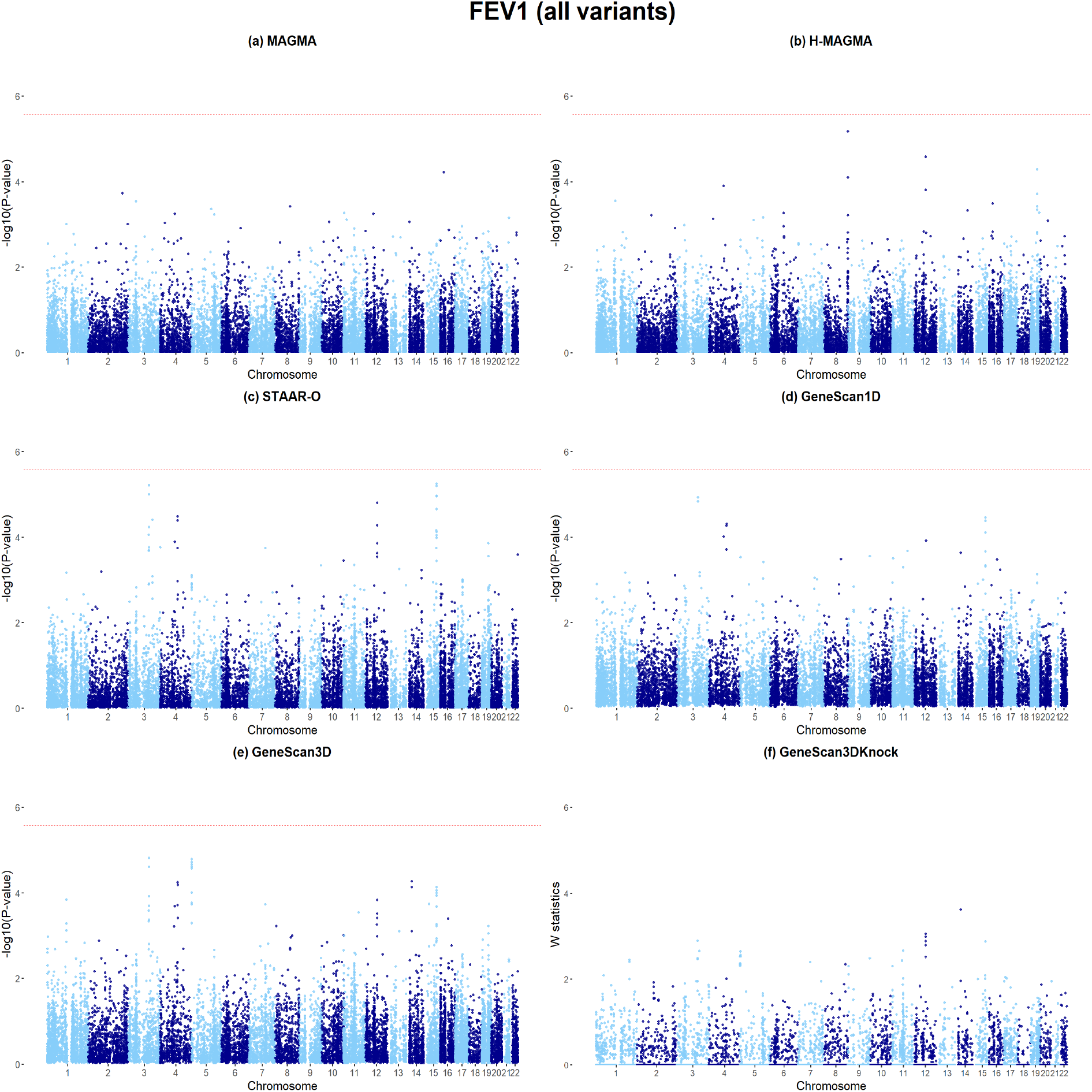
Applications to COPDGene whole-genome sequencing data (trait FEV1), for all variants. (a)-(f) Manhattan plots of MAGMA, H-MAGMA, STAAR-O, GeneScan1D, GeneScan3D and GeneScan3DKnock results, respectively.

**Figure S8:**
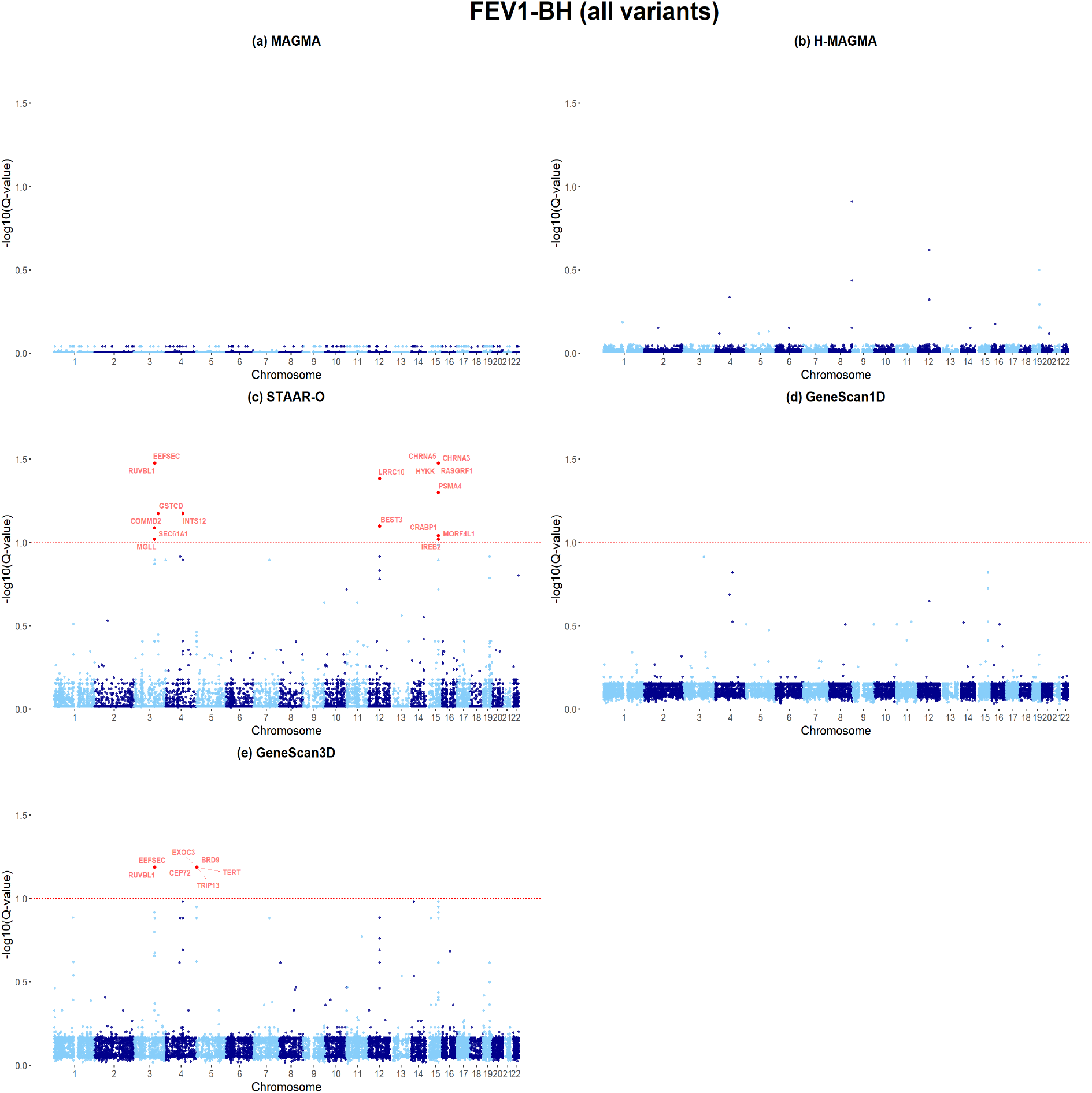
Applications to COPDGene whole-genome sequencing data (trait FEV1), for all variants. (a)-(e) Manhattan plots of MAGMA, H-MAGMA, STAAR-O, GeneScan1D and GeneScan3D and GeneScan3DKnock results using BH for FDR control, respectively.

**Figure S9:**
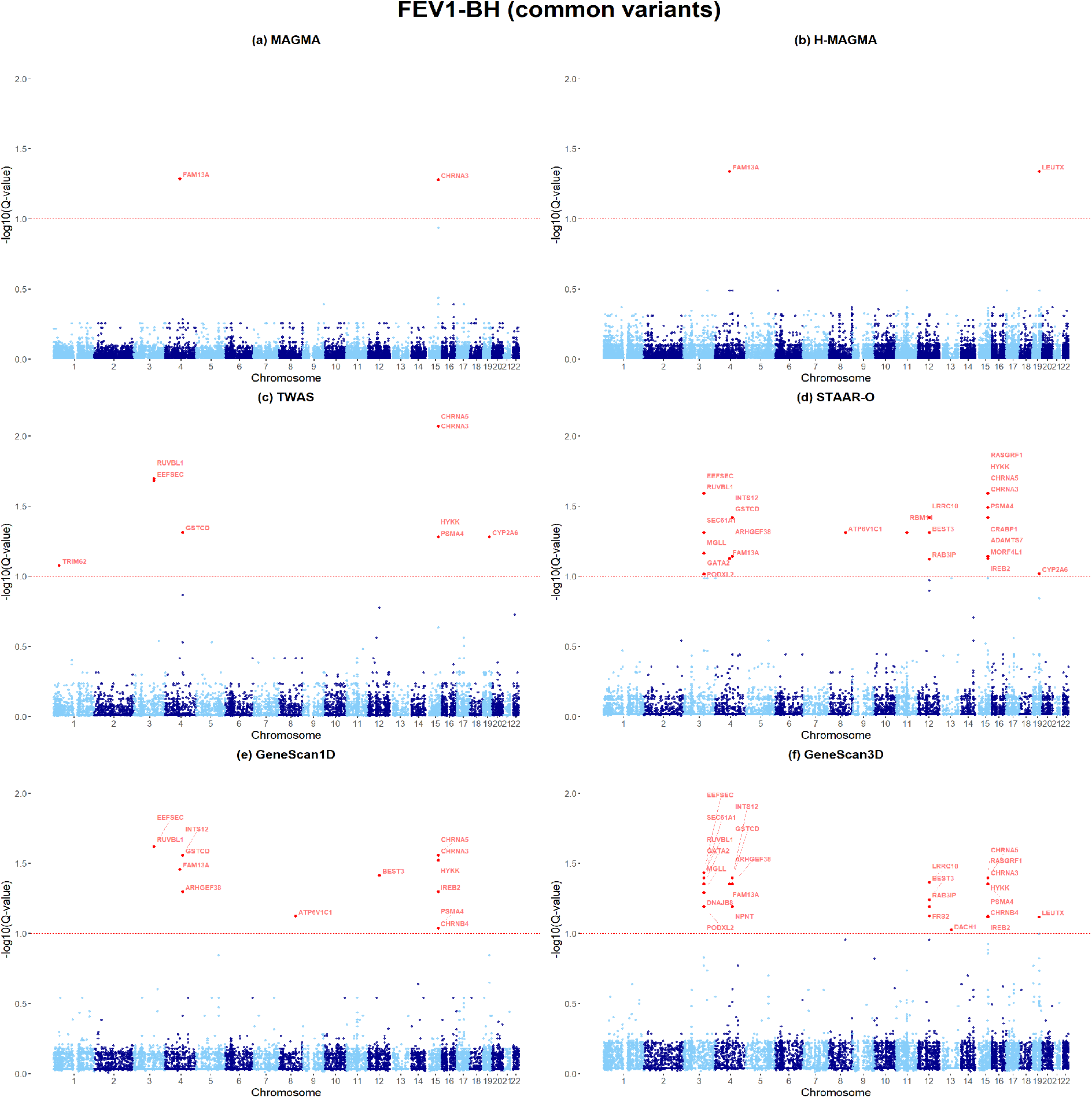
Applications to COPDGene whole-genome sequencing data (trait FEV1), for common variants. (a)-(f) Manhattan plots of MAGMA, H-MAGMA, TWAS, STAAR-O, GeneScan1D and GeneScan3D and GeneScan3DKnock results using BH for FDR control, respectively.

**Figure S10:**
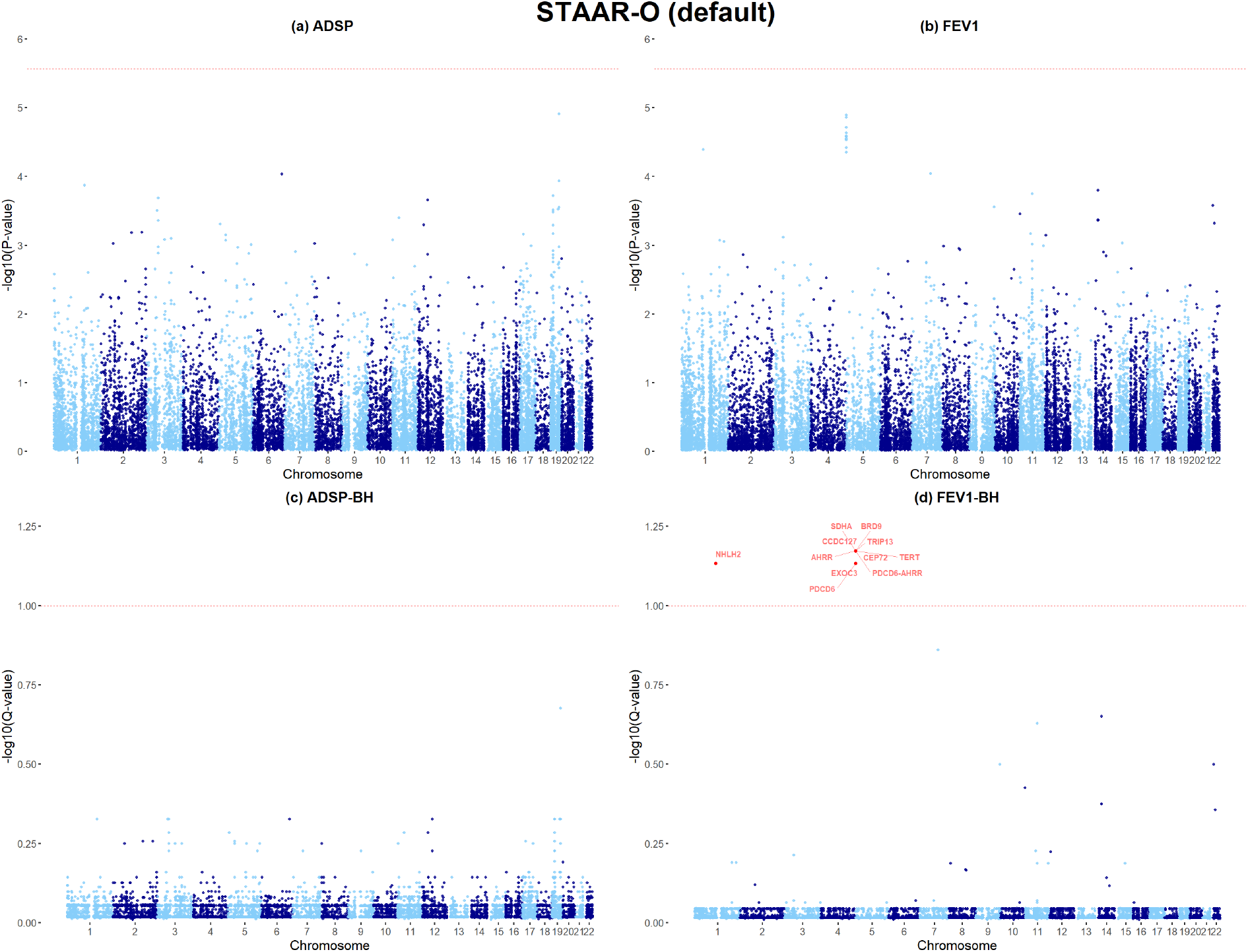
Applications to ADSP and COPDGene (trait FEV1) whole-genome sequencing data for STAAR-O default settings. (a)-(b) Manhattan plots of ADSP and FEV1 using Bonferroni threshold, respectively. (c)-(d) Manhattan plots of ADSP and FEV1 results BH for FDR control, respectively.

**Figure S11:**
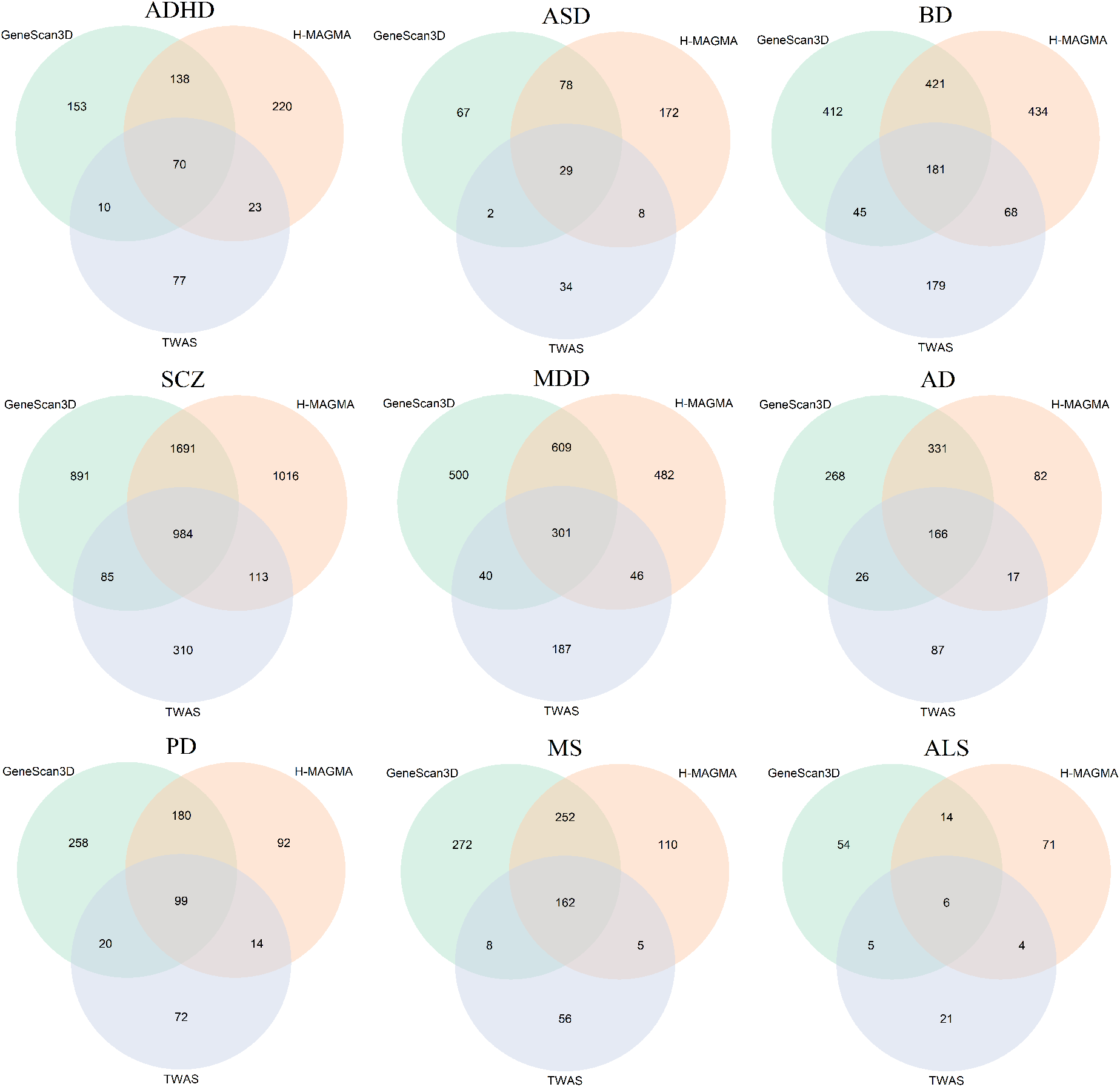
Overlap between significant genes of GeneScan3D, H-MAGMA and TWAS for 9 neuropsychiatric and neurodegenerative traits.

**Figure S12:**
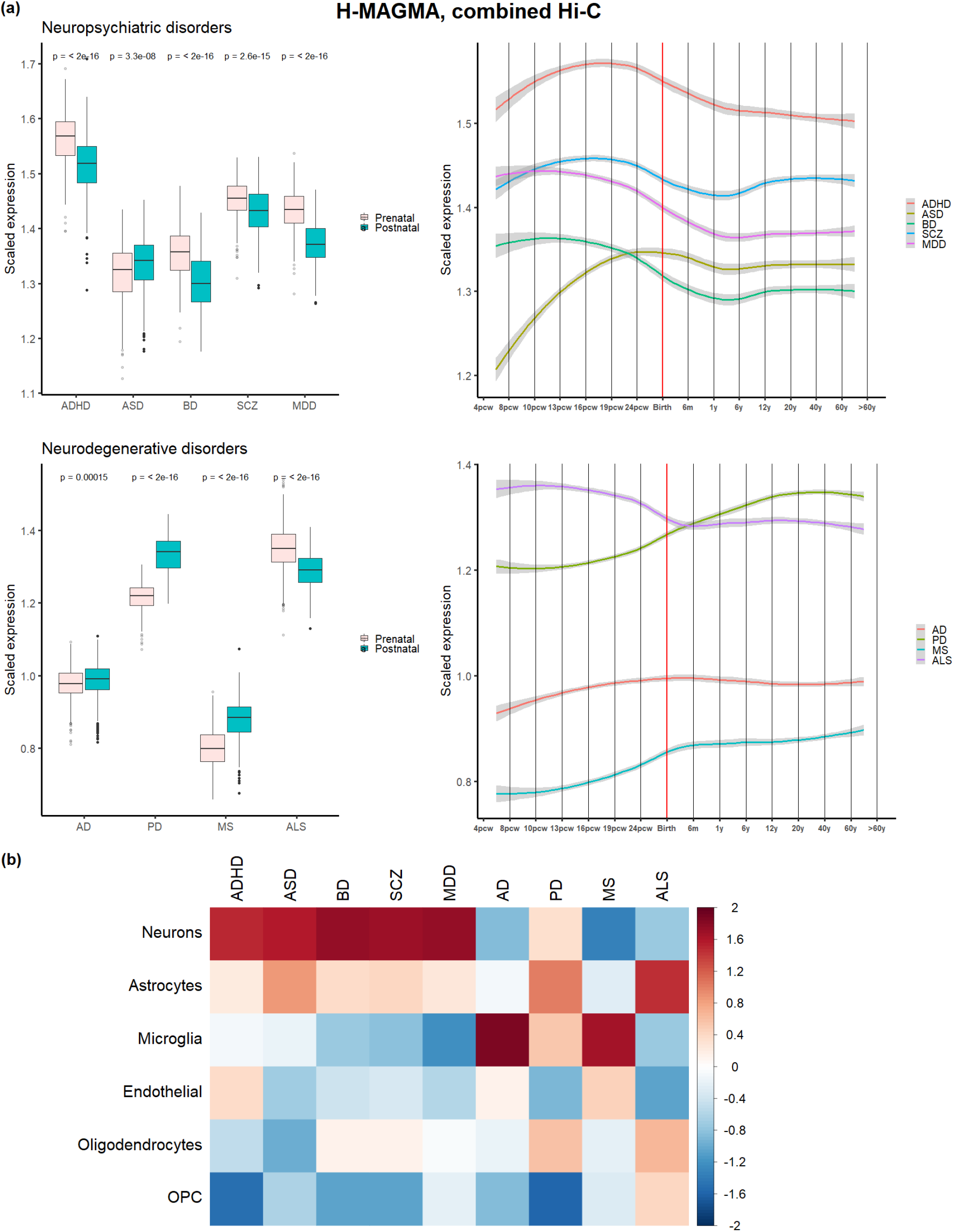
**(a) Human brain developmental expression of H-MAGMA significant genes for each brain-disorder**, combined Hi-C for Adult brain, Fetal brain GZ and CP layers. Pvalues of Wilcoxon rank sum tests are shown in the boxplots to compare independent prenatal and postnatal samples. **(b) Cell-type expression profiles of H-MAGMA significant genes**.

**Figure S13:**
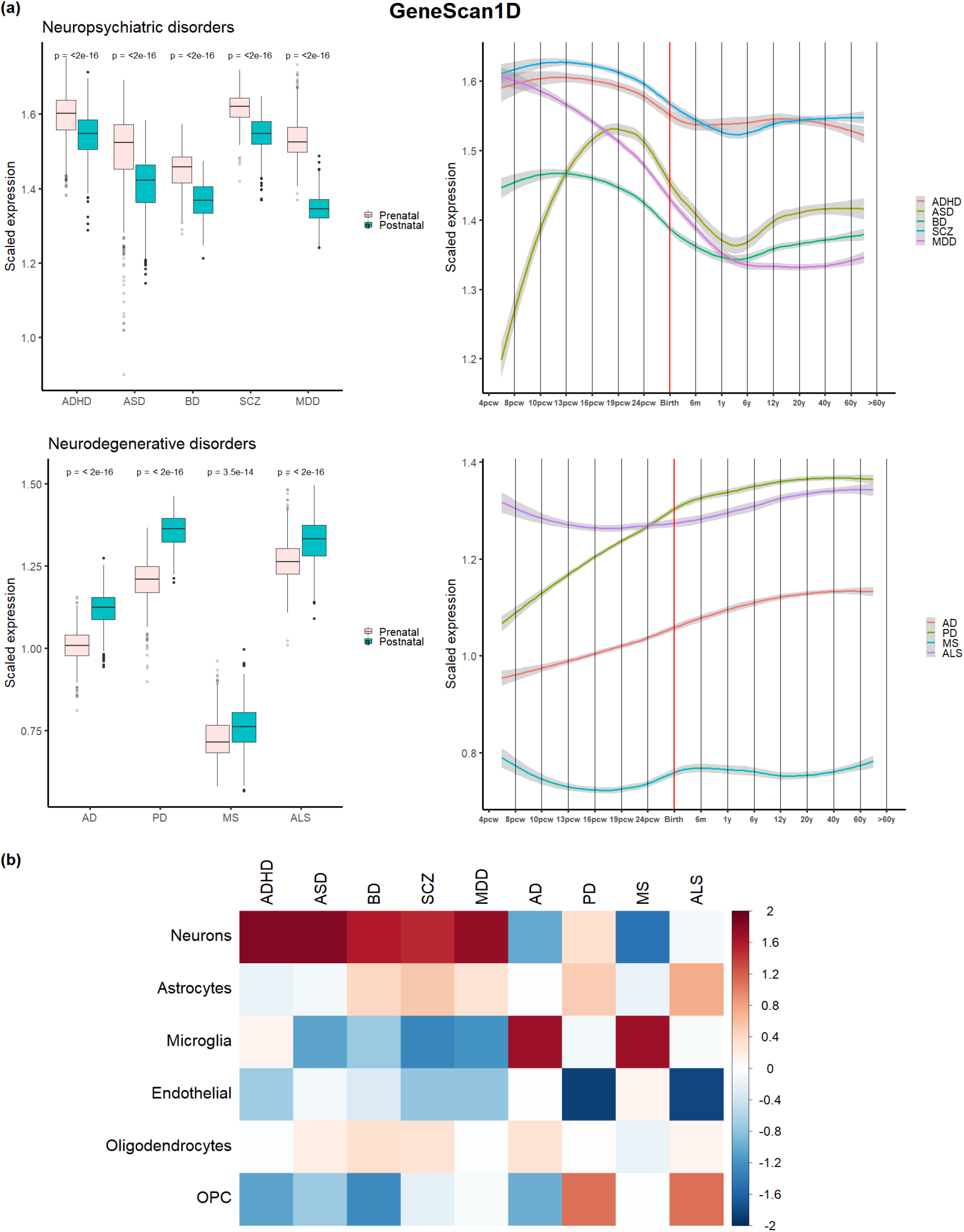
**(a) Human brain developmental expression of GeneScan1D significant genes for each brain-disorder**. P-values of Wilcoxon rank sum tests are shown in the boxplots to compare independent prenatal and postnatal samples. **(b) Cell-type expression profiles of GeneScan1D significant genes**.

**Figure S14:**
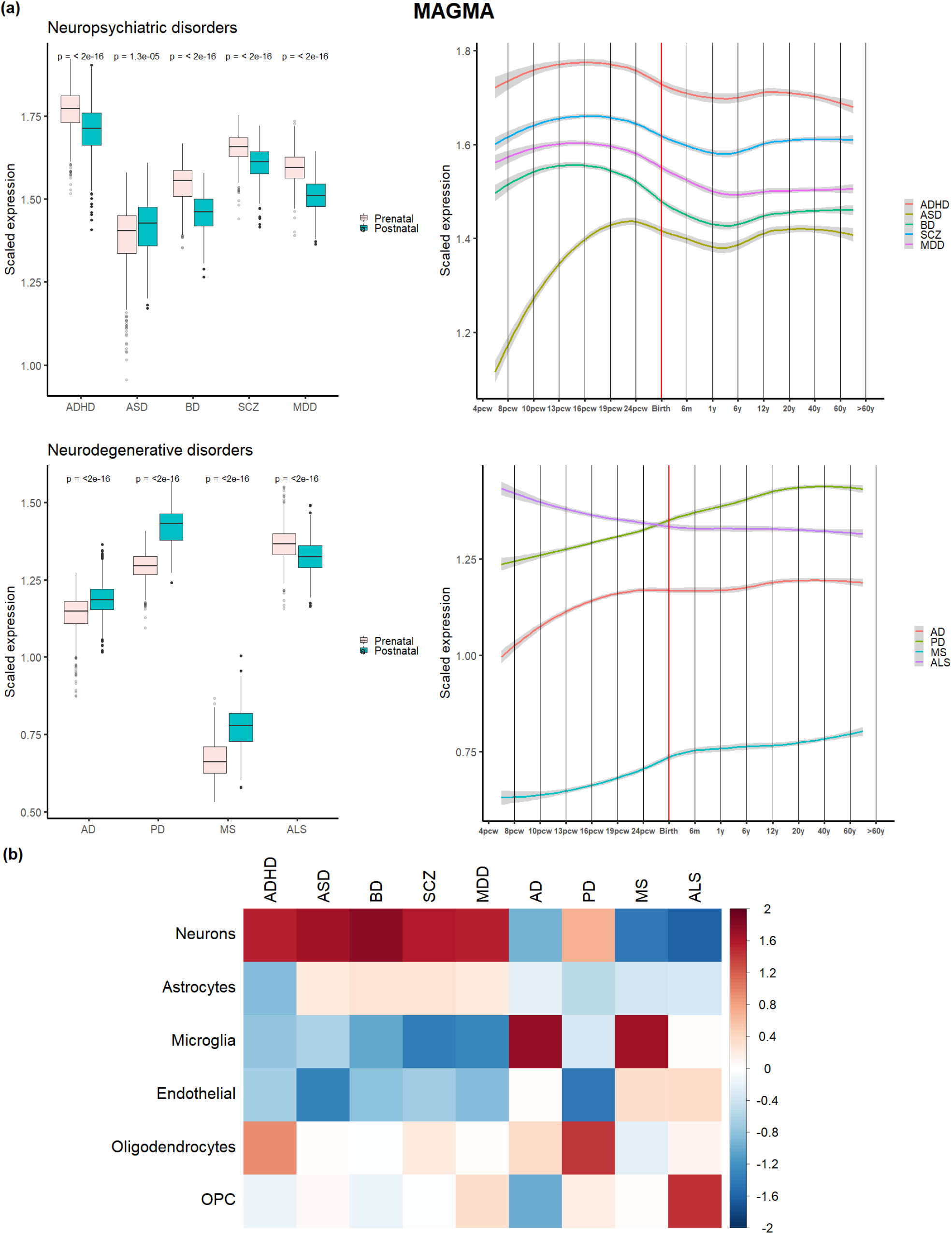
**(a) Human brain developmental expression of MAGMA significant genes for each brain-disorder**. P-values of Wilcoxon rank sum tests are shown in the boxplots to compare independent prenatal and postnatal samples. **(b) Cell-type expression profiles of MAGMA significant genes**.

**Figure S15:**
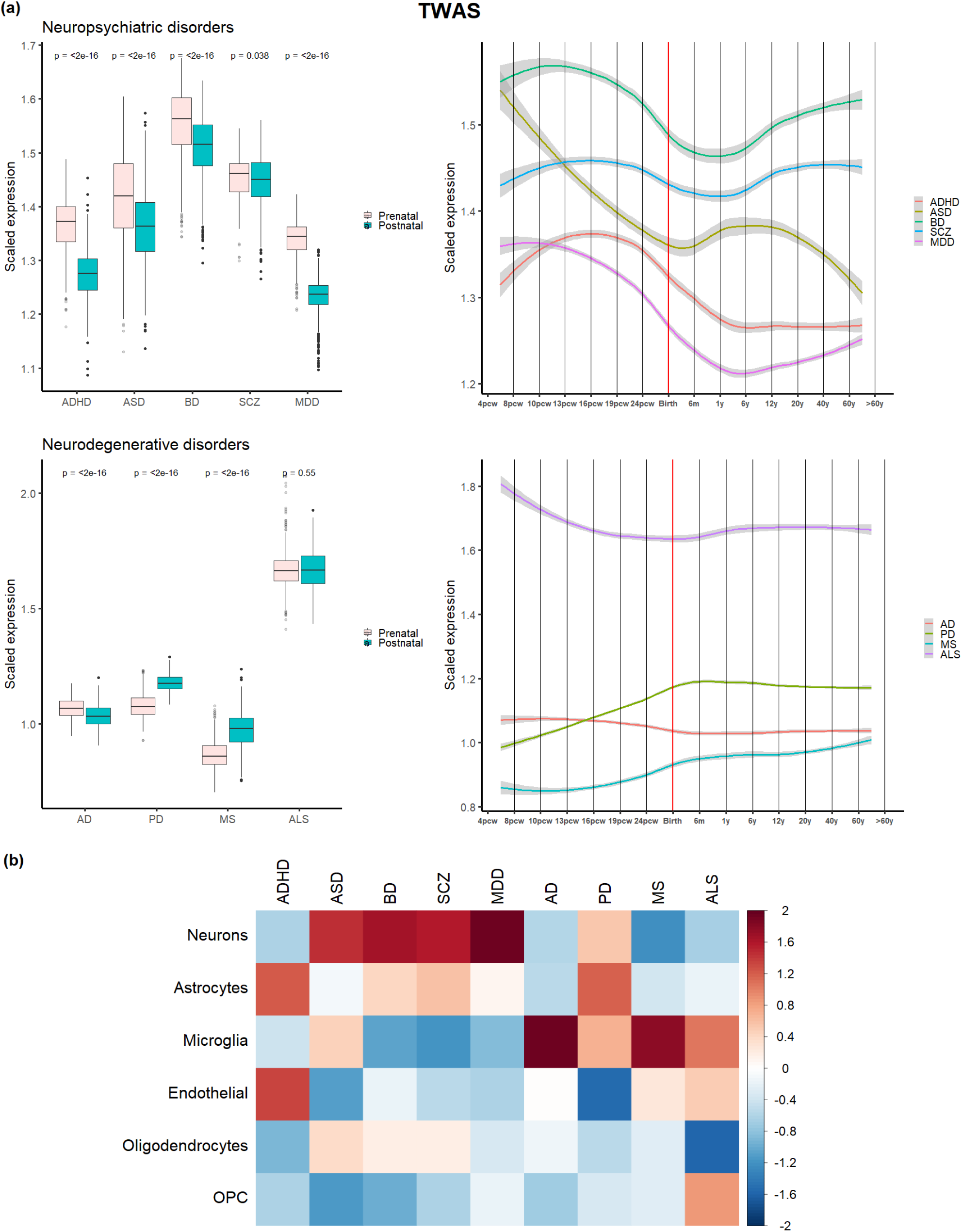
**(a) Human brain developmental expression of TWAS significant genes for each brain-disorder**. P-values of Wilcoxon rank sum tests are shown in the boxplots to compare independent prenatal and postnatal samples. **(b) Cell-type expression profiles of TWAS significant genes**.

**Table S1:** Type-I error rate of the GeneScan1D and GeneScan3D tests. Results are shown for tests based on all variants and common variants only, for continuous and binary traits.

| Trait | Significance level | GeneScan1D | GeneScan3D<br>($R = 2$ enhancers) |
| --- | --- | --- | --- |
| Common variants |  |  |  |
| Binary | $10^{-3}$ | 0.0011 | 0.0010 |
| | $10^{-4}$ | 9.67E-05 | 9.89E-05 |
| | $10^{-5}$ | 1.04E-05 | 1.04E-05 |
| | $2.5 \times 10^{-6}$ | 3.00E-06 | 2.50E-06 |
| Continuous | $10^{-3}$ | 0.0011 | 0.0011 |
| | $10^{-4}$ | 1.06E-04 | 1.03E-04 |
| | $10^{-5}$ | 1.02E-05 | 1.03E-05 |
| | $2.5 \times 10^{-6}$ | 2.90E-06 | 3.10E-06 |
| All variants |  |  |  |
| Binary | $10^{-3}$ | 8.21E-04 | 8.45E-04 |
| | $10^{-4}$ | 7.48E-05 | 7.69E-05 |
| | $10^{-5}$ | 7.50E-06 | 6.40E-06 |
| | $2.5 \times 10^{-6}$ | 2.50E-06 | 1.70E-06 |
| Continuous | $10^{-3}$ | 0.0011 | 0.0011 |
| | $10^{-4}$ | 1.02E-04 | 1.02E-04 |
| | $10^{-5}$ | 1.04E-05 | 1.03E-05 |
| | $2.5 \times 10^{-6}$ | 2.70E-06 | 2.90E-06 |

**Table S2:**
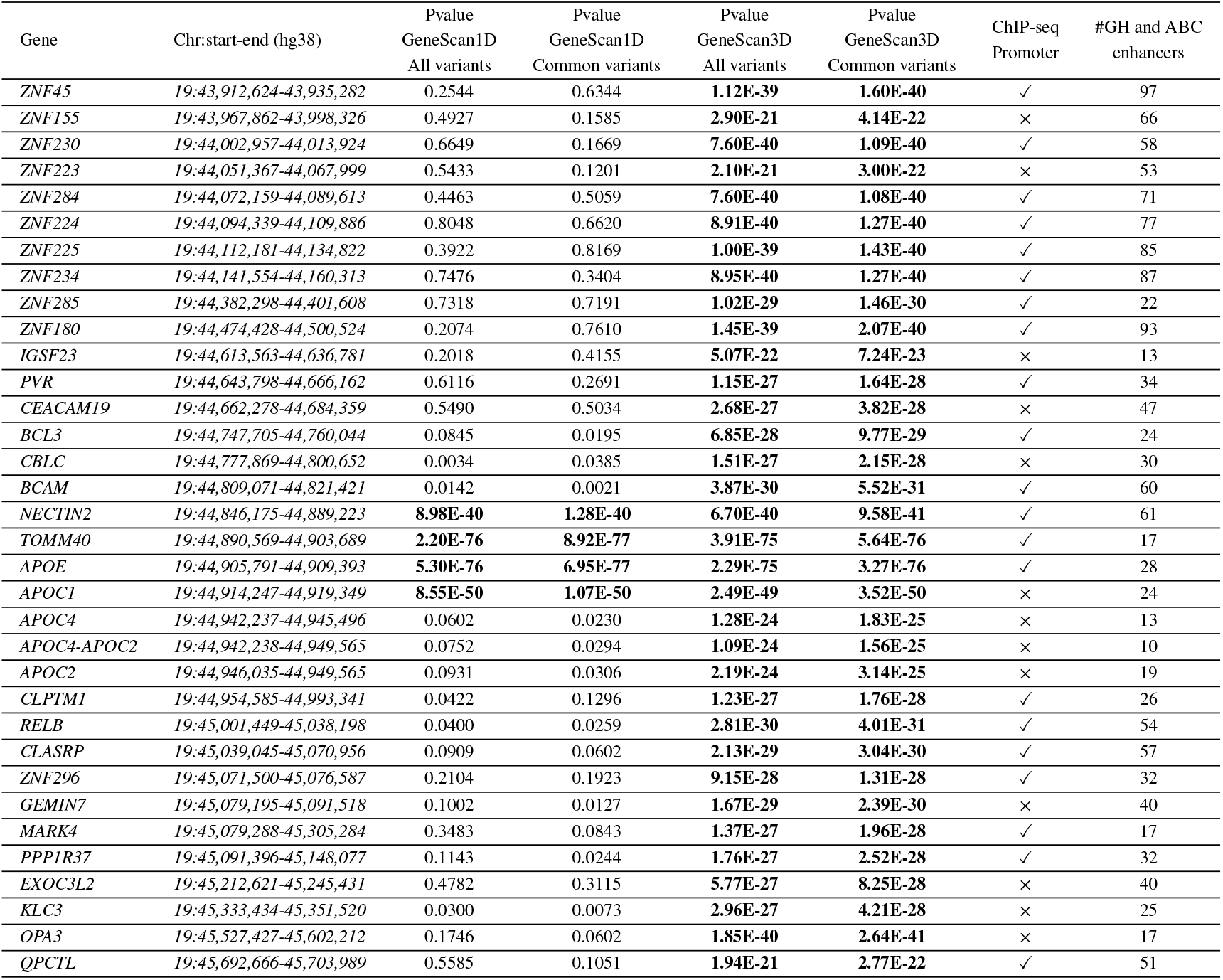
GeneScan1D/3D results for ADSP data. For all variants and/or common variants, genes significantly associated with AD in at least one of the GeneScan1D/3D analyses are reported. The p-values smaller than 2.7 × 10^−6^ are shown in boldface font.

**Table S3:** Windows with minimum p-value for the genes significant in the GeneScan3D analyses (common variants only) are shown (AD). For each significant gene, the minimum p-value for the 1-D windows is reported, along with the best 1-D window spanning the gene plus buffer region. The minimum p-value for the 3-D windows along with the corresponding regulatory element, and the strength of the enhancer-gene link are also reported.

| Gene | min-p<br>1-D windows | Best 1-D window (hg38) | min-p<br>3-D windows | Best RE with enhancer score (hg38) |
| --- | --- | --- | --- | --- |
| *ZNF45* | 0.1043 | *chr19:43,931,258-43,932,101* | 1.20E-42 | GH19F044903: *chr19:44,903,704-44,907,334*<br>GH link score 18.49 |
| *ZNF155* | 0.0084 | *chr19:43,994,962-43,995,935* | 4.26E-24 | GH19F044923: *chr19:44,923,872-44,928,016*<br>GH link score 21.11 |
| *ZNF230* | 0.0097 | *chr19:44,009,944-44,010,922* | 1.43E-42 | GH19F044903: *chr19:44,903,704-44,907,334*<br>GH link score 13.18 |
| *ZNF223* | 0.0203 | *chr19:44,059,376-44,060,351* | 3.99E-24 | GH19F044923: *chr19:44,923,872-44,928,016*<br>GH link score 12.64 |
| *ZNF284* | 0.0448 | *chr19:44,069,650-44,070,618* | 1.13E-42 | GH19F044903: *chr19:44,903,704-44,907,334*<br>GH link score 15.86 |
| *ZNF224* | 0.0608 | *chr19:44,091,826-44,092,807* | 1.13E-42 | GH19F044903: *chr19:44,903,704-44,907,334*<br>GH link score 20.39 |
| *ZNF225* | 0.0269 | *chr19:44,139,241-44,139,818* | 1.28E-42 | GH19F044903: *chr19:44,903,704-44,907,334*<br>GH link score 12.72 |
| *ZNF234* | 0.0224 | *chr19:44,142,512-44,143,203* | 1.13E-42 | GH19F044903: *chr19:44,903,704-44,907,334*<br>GH link score 12.18 |
| *ZNF285* | 0.0765 | *chr19:44,389,350-44,390,328* | 4.66E-32 | GH19F044889: *chr19:44,889,191-44,894,261*<br>GH link score 1.21 |
| *ZNF180* | 0.1769 | *chr19:44,474,408-44,475,380* | 1.58E-42 | GH19F044903: *chr19:44,903,704-44,907,334*<br>GH link score 18.84 |
| *IGSF23* | 0.0357 | *chr19:44,608,576-44,609,451* | 3.99E-24 | GH19F044923: *chr19:44,923,872-44,928,016*<br>GH link score 7.95 |
| *PVR* | 0.0083 | *chr19:44,661,414-44,666,407* | 7.02E-30 | ABC enhancer: *chr19:44,890,614-44,891,469*<br>ABC score 0.0379 |
| *CEACAM19* | 0.0147 | *chr19:44,663,283-44,664,277* | 4.68E-30 | ABC enhancer: *chr19:44,890,614-44,891,469*<br>ABC score 0.0366 |
| *BCL3* | 0.0013 | *chr19:44,758,712-44,759,652* | 4.68E-30 | ABC enhancer: *chr19:44,890,614-44,891,469*<br>ABC score 0.0411 |
| *CBLC* | 0.0019 | *chr19:44,783,946-44,784,890* | 4.68E-30 | ABC enhancer: *chr19:44,890,614-44,891,469*<br>ABC score 0.076 |
| *BCAM* | 7.28E-05 | *chr19:44,820,132-44,821,030* | 6.97E-33 | GH19F044909: *chr19:44,909,534-44,915,843*<br>GH link score 4.9 |
| *NECTIN2* | 1.20E-42 | *chr19:44,888,835-44,889,771* | 1.25E-42 | GH19F044903: *chr19:44,903,704-44,907,334*<br>GH link score 5.79 |
| *TOMM40* | 3.37E-78 | *chr19:44,908,102-44,908,684* | 1.25E-77 | promoter |
| *APOE* | 3.37E-78 | *chr19:44,907,807-44,908,799* | 8.36E-78 | promoter |
| *APOC1* | 3.92E-52 | *chr19:44,918,285-44,919,189* | 3.92E-52 | ABC enhancer: *chr19:44,906,123-44,906,623*<br>ABC score 0.0334 |
| *APOC4* | 0.0018 | *chr19:44,943,215-44,944,186* | 7.88E-27 | ABC enhancer: *chr19:44,912,770-44,913,270*<br>ABC score 0.0302 |
| *APOC4-APOC2* | 0.0018 | *chr19:44,943,215-44,944,186* | 7.88E-27 | ABC enhancer: *chr19:44,912,770-44,913,270*<br>ABC score 0.0302 |
| *APOC2* | 0.0018 | *chr19:44,943,144-44,944,134* | 7.88E-27 | ABC enhancer: *chr19:44,912,770-44,913,270*<br>ABC score 0.0302 |
| *CLPTM1* | 0.0117 | *chr19:44,967,087-44,968,058* | 1.87E-29 | ABC enhancer: *chr19:44,890,614-44,891,469*<br>ABC score 0.0737 |
| *RELB* | 0.0004 | *chr19:45,020,477-45,021,421* | 6.67E-33 | GH19F044909: *chr19:44,909,534-44,915,843*<br>GH link score 11.26 |
| *CLASRP* | 0.0012 | *chr19:45,063,100-45,064,084* | 4.45E-32 | GH19F044889: *chr19:44,889,191-44,894,261*<br>GH link score 1.55 |
| *ZNF296* | 0.0349 | *chr19:45,076,520-45,081,506* | 7.02E-30 | ABC enhancer: *chr19:44,890,614-44,891,469*<br>ABC score 0.0202 |
| *GEMIN7* | 0.0008 | *chr19:45,084,166-45,089,162* | 4.66E-32 | GH19F044889: *chr19:44,889,191-44,894,261*<br>GH link score 1.9 |
| *MARK4* | 0.0008 | *chr19:45,084,166-45,089,162* | 9.36E-30 | ABC enhancer: *chr19:44,890,282-44,891,469*<br>ABC score 0.0301 |
| *PPP1R37* | 0.0002 | *chr19:45,086,407-45,091,400* | 9.36E-30 | ABC enhancer: *chr19:44,890,284-44,891,469*<br>ABC score 0.0262 |
| *EXOC3L2* | 0.0184 | *chr19:45,211,680-45,212,582* | 7.02E-30 | ABC enhancer: *chr19:44,890,308-44,891,469*<br>ABC score 0.0263 |
| *KLC3* | 0.0005 | *chr19:45,332,046-45,333,045* | 7.02E-30 | ABC enhancer: *chr19:44,890,282-44,891,469*<br>ABC score 0.0322 |
| *OPA3* | 0.0010 | *chr19:45,592,914-45,593,889* | 1.13E-42 | GH19F044903: *chr19:44,903,704-44,907,334*<br>GH link score 4.48 |
| *QPCTL* | 0.0116 | *chr19:45,691,716-45,692,686* | 3.99E-24 | GH19F044923: *chr19:44,923,872-44,928,016*<br>GH link score 12.14 |

**Table S4:** GeneScan3DKnock results for ADSP data. For all variants and/or common variants, genes significantly associated with AD for the GeneScan3DKnock test at FDR 10% are shown. ‘-’ represents genes that are not significant under all variants or common variants (with q-value> FDR 10%).

| Gene | Chr:start-end (hg38) | W statistic<br>All variants | q-value<br>All variants | W statistic<br>Common variants | q-value<br>Common variants |
| --- | --- | --- | --- | --- | --- |
| *PPP1R17* | *chr7:31,687,215-31,708,455* | - | - | 2.70 | 0.0957 |
| *HIPK3* | *chr11:33,256,672-33,357,023* | 3.20 | 0.0400 | - | - |
| *NECTIN1* | *chr11:119,623,408-119,729,200* | - | - | 3.23 | 0.0200 |
| *ZNF646* | *chr16:31,074,422-31,084,196* | - | - | 2.71 | 0.0957 |
| *ZNF843* | *chr16:31,432,593-31,443,160* | - | - | 2.77 | 0.0957 |
| *ZNF45* | *chr19:43,912,624-43,935,282* | 3.74 | 0.0211 | 3.74 | 0.0105 |
| *ZNF155* | *chr19:43,967,862-43,998,326* | 8.82 | 0.0118 | 8.82 | 0.0105 |
| *ZNF230* | *chr19:44,002,957-44,013,924* | 4.10 | 0.0118 | 4.10 | 0.0105 |
| *ZNF223* | *chr19:44,051,367-44,067,999* | 8.82 | 0.0118 | 8.82 | 0.0105 |
| *ZNF284* | *chr19:44,072,159-44,089,613* | 4.41 | 0.0118 | 4.41 | 0.0105 |
| *ZNF224* | *chr19:44,094,339-44,109,886* | 4.41 | 0.0118 | 4.41 | 0.0105 |
| *ZNF225* | *chr19:44,112,181-44,134,822* | 4.41 | 0.0118 | 4.41 | 0.0105 |
| *ZNF234* | *chr19:44,141,554-44,160,313* | 4.10 | 0.0118 | 4.10 | 0.0105 |
| *ZNF180* | *chr19:44,474,428-44,500,524* | 3.74 | 0.0211 | 3.74 | 0.0105 |
| *IGSF23* | *chr19:44,613,563-44,636,781* | 8.82 | 0.0118 | 8.82 | 0.0105 |
| *BCAM* | *chr19:44,809,071-44,821,421* | 9.06 | 0.0118 | 9.06 | 0.0105 |
| *NECTIN2* | *chr19:44,846,175-44,889,223* | 8.07 | 0.0118 | 8.07 | 0.0105 |
| *TOMM40* | *chr19:44,890,569-44,903,689* | 10.98 | 0.0118 | 10.98 | 0.0105 |
| *APOE* | *chr19:44,905,791-44,909,393* | 10.98 | 0.0118 | 10.98 | 0.0105 |
| *APOC1* | *chr19:44,914,247-44,919,349* | 5.20 | 0.0118 | 5.20 | 0.0105 |
| *CLPTM1* | *chr19:44,954,585-44,993,341* | 4.09 | 0.0118 | 4.09 | 0.0105 |
| *RELB* | *chr19:45,001,449-45,038,198* | 9.05 | 0.0118 | 9.05 | 0.0105 |
| *OPA3* | *chr19:45,527,427-45,602,212* | 4.41 | 0.0118 | 4.41 | 0.0105 |
| *QPCTL* | *chr19:45,692,666-45,703,989* | 8.82 | 0.0118 | 8.82 | 0.0105 |

**Table S5:** Replication results for ADSP data. The list includes all genes that have been identified as significant across all the different tests/analyses (including BH-adjusted tests). Genes identified by GeneScan3DKnock are shown in italic font. The p-values smaller than 0.05 are shown in boldface font.

| Gene | Pvalue GeneScan3D | Pvalue GeneScan1D |
| --- | --- | --- |
| CHI3L2 | 6.14E-01 | 3.27E-01 |
| NOS1AP | 8.32E-01 | 1.56E-01 |
| PDE12 | **1.47E-05** | **4.88E-06** |
| PLK4 | 6.10E-01 | 6.73E-01 |
| TENT4A | 8.52E-02 | 6.36E-02 |
| RIOK1 | 2.79E-01 | 3.99E-01 |
| ADGRG6 | 7.33E-01 | 7.33E-01 |
| IPCEF1 | 5.93E-01 | 2.03E-01 |
| *PPP1R17* | **4.96E-02** | 5.56E-02 |
| INIP | **6.40E-03** | **1.93E-02** |
| SLC46A2 | **7.38E-03** | **5.94E-03** |
| ZFP37 | 7.40E-02 | 7.94E-01 |
| *HIPK3* | 1.81E-01 | 1.50E-01 |
| *NECTIN1* | 4.23E-01 | 4.23E-01 |
| ELF1 | 4.38E-01 | 5.91E-01 |
| KBTBD7 | 3.33E-01 | 5.41E-01 |
| RBPMS2 | 1.65E-01 | 8.64E-01 |
| *ZNF646* | **2.52E-05** | **3.68E-05** |
| BCKDK | **2.45E-06** | **9.07E-07** |
| PRSS8 | **5.04E-05** | **3.92E-05** |
| FUS | **3.01E-05** | **1.73E-02** |
| *ZNF843* | **4.10E-04** | 3.36E-01 |
| TRIR | 4.01E-01 | 5.79E-01 |
| *ZNF45* | **0.00E+00** | **3.08E-02** |
| *ZNF155* | **0.00E+00** | 6.35E-02 |
| *ZNF230* | **0.00E+00** | **3.50E-03** |
| *ZNF223* | **0.00E+00** | **3.87E-03** |
| *ZNF284* | **0.00E+00** | 5.10E-02 |
| *ZNF224* | **0.00E+00** | 1.81E-01 |
| *ZNF225* | **0.00E+00** | 1.86E-01 |
| *ZNF234* | **0.00E+00** | **7.73E-05** |
| *ZNF285* | **0.00E+00** | **2.19E-02** |
| *ZNF180* | **0.00E+00** | **5.97E-03** |
| *IGSF23* | **0.00E+00** | **1.53E-14** |
| PVR | **0.00E+00** | **8.50E-24** |
| CEACAM19 | **0.00E+00** | **9.97E-24** |
| BCL3 | **0.00E+00** | **5.23E-56** |
| CBLC | **0.00E+00** | **1.05E-39** |
| *BCAM* | **0.00E+00** | **1.07E-142** |
| *NECTIN2* | **0.00E+00** | **0.00E+00** |
| *TOMM40* | **0.00E+00** | **0.00E+00** |
| *APOE* | **0.00E+00** | **0.00E+00** |
| *APOC1* | **0.00E+00** | **0.00E+00** |
| APOC4 | **0.00E+00** | **9.39E-33** |
| APOC4-APOC2 | **0.00E+00** | **9.39E-33** |
| APOC2 | **0.00E+00** | **5.82E-33** |
| *CLPTM1* | **0.00E+00** | **7.83E-48** |
| *RELB* | **0.00E+00** | **1.08E-44** |
| CLASRP | **0.00E+00** | **1.67E-27** |
| ZNF296 | **0.00E+00** | **6.51E-14** |
| GEMIN7 | **0.00E+00** | **3.37E-17** |
| MARK4 | **0.00E+00** | **4.24E-36** |
| PPP1R37 | **0.00E+00** | **1.20E-36** |
| EXOC3L2 | **0.00E+00** | **1.55E-20** |
| KLC3 | **0.00E+00** | **3.27E-06** |
| *OPA3* | **0.00E+00** | **3.79E-04** |
| *QPCTL* | **0.00E+00** | **2.44E-03** |
| LCA5L | 6.89E-01 | 8.15E-01 |

**Table S6:**
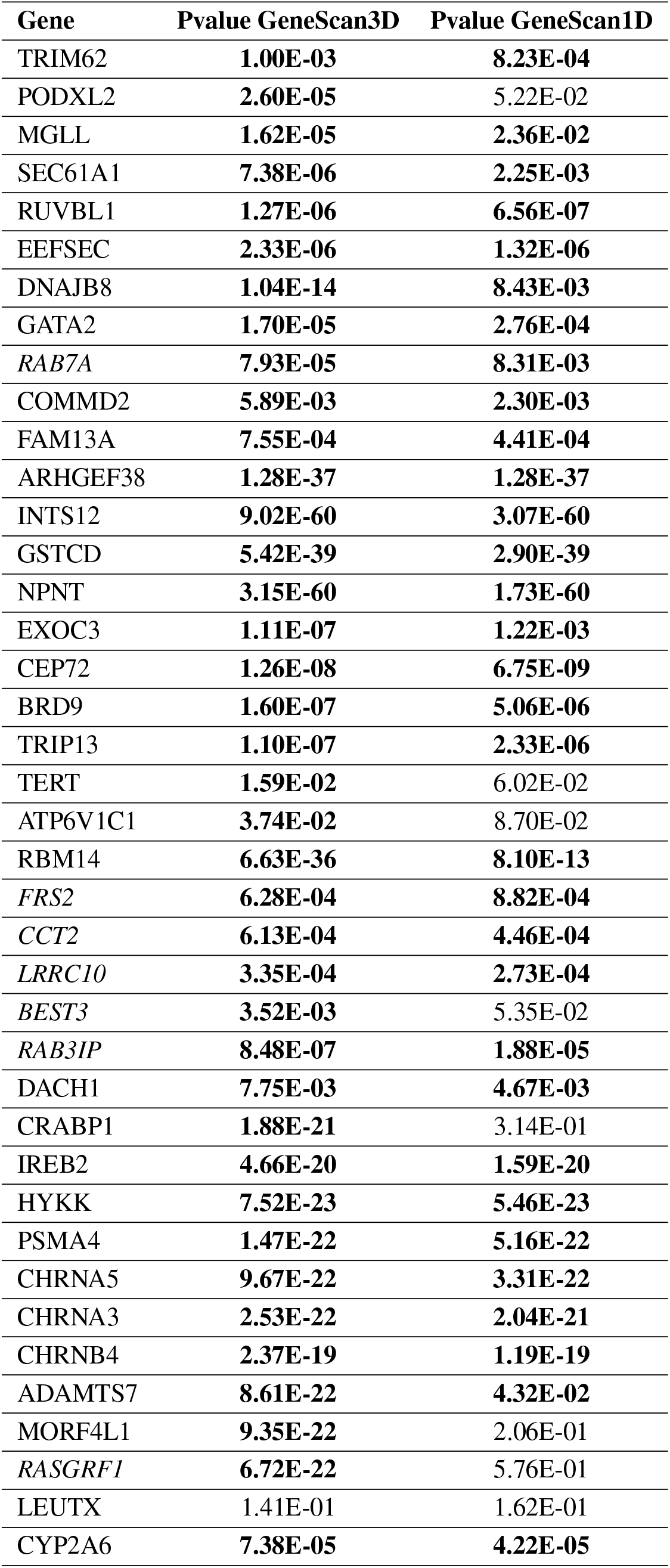
Replication results for COPDGene data (trait FEV1). The list includes all genes that have been identified as significant across all the different tests/analyses (including BH-adjusted tests). Genes identified by GeneScan3DKnock are shown in italic font. The p-values smaller than 0.05 are shown in boldface font.

**Table S7:** The number of significant genes (*p* ≤ 10^−3^) associated with each brain disorder for GeneScan tests, MAGMA/H-MAGMA and TWAS.

| Disorder | GeneScan1D | GeneScan3D<br>combined Hi-C | MAGMA | H-MAGMA<br>combined Hi-C | TWAS |
| --- | --- | --- | --- | --- | --- |
| ADHD | 193 | 371 | 206 | 451 | 180 |
| ASD | 95 | 176 | 101 | 287 | 73 |
| BD | 421 | 1059 | 475 | 1104 | 474 |
| SCZ | 1742 | 3651 | 1753 | 3804 | 1496 |
| MDD | 657 | 1450 | 665 | 1438 | 575 |
| AD | 308 | 791 | 246 | 596 | 296 |
| PD | 214 | 557 | 171 | 385 | 205 |
| MS | 364 | 694 | 338 | 529 | 231 |
| ALS | 55 | 79 | 51 | 95 | 36 |

